# Explainable Longitudinal Machine Learning for Dementia Progression Using Cognitive and MRI Biomarkers

**DOI:** 10.64898/2026.07.12.26357878

**Authors:** Gifty Duah, Eric Nyarko, Justice Yaw Effah, Isaac Boateng Numoah, Anani Lotsi

## Abstract

Dementia is a progressive neurological condition characterized by cognitive decline and structural brain changes that evolve. Longitudinal modeling of these changes is important for improving disease monitoring, identifying progression patterns, and supporting early risk stratification. This study developed an explainable longitudinal machine-learning framework for dementia progression, using cognitive and Magnetic Resonance Imaging (MRI)-derived biomarkers from the Open Access Series of Imaging Studies (OASIS-2) longitudinal dataset. The dataset included 150 subjects and 373 repeated observations classified as Non-demented, Demented, or Converted. Current-visit features, previous-visit features, and slope-based temporal features were constructed from Mini-Mental State Examination, Clinical Dementia Rating, normalized whole-brain volume, estimated total intracranial volume, atlas scaling factor, Age, and MRI delay. Baseline models were compared with a longitudinal gradient-boosted model, using patient-level splitting to reduce data leakage across repeated visits. The proposed longiGradient Gradient boosting model achieved the best held-out test performance, with an accuracy of 88.16%, a macro F1-score of 0.776, and a weighted F1-score of 0.860. The model showed strong classification performance for Demented and Non-demented individuals, while converted cases remained more difficult to identify. A regularized gradient boosting model was also evaluated as an overfitting sensitivity analysis; although it reduced the perfect training fit, it did not improve held-out test performance. Feature importance, permutation importance, and SHapley Additive exPlanations identified Clinical Dementia Rating as the dominant predictor, with slope-based Clinical Dementia Rating providing additional longitudinal information. These findings suggest that combining cognitive measures, MRI-derived biomarkers, and temporal feature engineering can improve dementia progression modeling, although external validation in larger longitudinal cohorts is needed.

## 1. INTRODUCTION

Alzheimer’s disease and related dementias represent a growing global health challenge, characterized by progressive cognitive decline and neurodegeneration that varies considerably across individuals [1–3]. By 2050, the number of people living with dementia worldwide is projected to nearly triple, reaching approximately 139 million [4]. Understanding the complex trajectories of cognitive deterioration and their relationship to structural brain changes is essential for early detection, disease monitoring, and the development of targeted interventions [5, 6]. The annual global economic cost of dementia exceeded USD 1 trillion in 2018 and continues to rise [7]. However, the progression of dementia is marked by significant heterogeneity in both timing and severity of symptoms, challenging conventional approaches to disease characterization and prediction [1, 8].

Traditional diagnostic approaches rely on cross-sectional data, which provide only a snapshot of an individual’s condition and fails to capture the dynamic nature of the disease progression [9]. Single-visit assessments are sensitive to individual variability, measurement noise, and floor or ceiling effects in cognitive scales [10, 11], and cannot differentiate stable from transitional cognitive states — an important limitation given the heterogeneous trajectory of mild cognitive impairment (MCI) [12, 13]. Longitudinal modeling approaches have therefore emerged as powerful tools for capturing the dynamic nature of dementia progression. Traditional linear mixed-effects models have provided foundational insights, with Moustafa et al. [14] demonstrating that Cognitive decline occurs across cognitively normal, MCI, and Alzheimer’s disease groups but at different rates. However, recent methodological advances have revealed the limitations of assuming linear progression, leading to more sophisticated approaches including semiparametric models [1], non-linear mixed-effects models [8], and disease progression models capable of capturing non-linear features of cognitive decline and neurodegeneration [15].

Central to understanding dementia progression is the relationship between structural brain changes and cognitive decline. Mounting evidence demonstrates that gray matter volume changes, particularly in temporal lobe structures, serve as robust predictors of cognitive deterioration [16–18]. Gavett et al. [19] identified a bifactor model of brain change that explained 59% of variance in cognitive decline, with global atrophy, temporolimbic, and medial temporal factors emerging as the strongest predictors. The Mini-Mental State Examination (MMSE) [10] and the Clinical Dementia Rating (CDR) [11] remains the most widely used clinical tool for staging dementia severity, while normalized whole brain volume (nWBV) provides a complementary structural biomarker that accounts for individual head-size differences and correlates with disease stage [17, 18]. The temporal dynamics of biomarker changes add further complexity: Lo et al. [20] demonstrated that trajectories of cerebrospinal fluid markers, glucose metabolism, and hippocampal volume vary across cognitive stages, supporting a hypothetical sequence wherein amyloid deposition precedes hypometabolism and hippocampal atrophy — a cascade formalized by [9] and corroborated by neuropathological staging evidence [21].

Recent methodological innovations have further enhanced the ability to model this complexity. Ribino et al. [22] introduced multivariate longitudinal clustering approaches that identify distinct subgroups based on co-occurring trajectory patterns, while Rakêt [8] developed models that explicitly separate disease progression from baseline cognitive capabilities, revealing a disease timeline spanning approximately 15 years from the earliest symptoms to severe dementia. Machine-learning (ML) approaches, including gradient-boosted ensemble methods and deep learning architectures, have demonstrated increasing utility for integrating multimodal and longitudinal clinical data [23–25]. SHAP (SHapley Additive exPlanations) [26, 27] has emerged as a principled framework for interpreting such models, providing patient-level feature attributions that are essential for clinical translation [28].

Despite these advances, significant challenges remain in accurately modeling the heterogeneous nature of dementia progression and translating longitudinal findings into clinically actionable insights. The present study addresses these challenges by applying a longitudinal ML framework to the Open Access Series of Imaging Studies (OASIS) dataset [29], incorporating temporal features derived from repeated cognitive assessments and structural Magnetic Resonance Imaging (MRI) measurements to classify and characterize Non-demented, demented, and converted individuals. By combining predictive modeling with SHAP-based interpretability, the study aims to identify the key biomarker trajectories driving dementia progression and to provide insights applicable to early risk stratification and clinical monitoring [5, 6, 24].

## 2. RELATED WORK

### 2.1. Longitudinal Modeling Approaches

The field has evolved from traditional linear mixed-effects models to more sophisticated ones approaches that better capture the non-linear and heterogeneous nature of dementia progression. Moustafa et al. [14] applied latent curve models to examine developmental trajectories across cognitively normal, MCI, and Alzheimer’s disease (AD) groups, finding that linear functional forms were adequate for clinical and neural measures, with cognitive decline occurring across all groups but at different rates; AD participants declined at approximately twice the rate of cognitively normal individuals on standardized cognitive composites. Gelir et al. [1] introduced semiparametric modeling using regression splines and mixed modeling techniques to capture non-linear AD progression-AlzheimAlzheimer’sAlzheimer’s Disease Assessment Scale-13 scores and ventricular volumes from the Alzheimer’s Disease Neuroimaging Initiative (ADNI) database, demonstrating significantly better fit than purely linear models (*p* < 0.001). Rakêt [8] developed non-linear mixed-effects disease progression models that explicitly represent disease stage, baseline cognition, and individual change trajectories as latent variables, estimating a cognitive decline timeline spanning approximately 15 years from the earliest subjective deficits to severe dementia; predicted disease time was significantly predictive of time since cognitive symptom onset (*p* < 0.0001), time since AD symptoms onset (*p* < 0.0001), and time since Alzheimer diagnosis (*p* < 0.0001), with Fluorodeoxyglucose Positron Emission Tomography (FDG-PET) explaining the most variation among biomarkers, followed by cerebrospinal fluid Aβ_1-42_/A*β*_1-40_ and florbetapir standardized Uptake Value ratio. Ribino et al. [22] proposed a multivariate time-series extension of *k*-means clustering to identify longitudinal subgroups compatible with Mild Behavioral Impairment preceding typical cognitive symptoms, achieving stable cluster solutions with silhouette coefficients exceeding 0.60.

### 2.2. Brain Volume Changes and Cognitive Decline

Consistent evidence demonstrates that gray matter volume changes, particularly in temporal regions, are strongly associated with cognitive deterioration. Fletcher et al. [16] examined 460 older adults and found that the global and temporal lobe gray matter volume change was the strongest predictor of cognitive change, with voxelwise analyses yielding *t*-values up to 13 in temporal lobe regions; a one standard deviation increase in the rate of gray matter atrophy was associated with an annual increase in cognitive decline rate of 0.057 SD. Gavett et al. [19] developed a bifactor model of brain change across 27 bilateral regions in 358 participants with up to eight annual visits, identifying global atrophy (β = 0.434), temporolimbic (β = 0.275), and medial temporal (β = 0.240) factors as the strongest predictors of cognitive slope, explaining 59*%* of variance in cognitive decline (Comparative Fit Index = 0.95, Tucker–Lewis Index = 0.924, Root Mean Square Error of Approximation = 0.065, Standardized Root Mean Square Residual = 0.037). Mungas et al. [30] showed in 120 subjects followed over three years that cortical gray matter atrophy predicted cognitive decline independently of subcortical lacune status (standardized *β* ≈0.40), while hippocampal atrophy predicted decline specifically in lacune-free participants. Chen et al. [31] subsequently confirmed that longitudinal changes in cortical thickness and hippocampal volume predict cognitive decline on standardized batteries, with Area Under the Curve (AUC) values of 0.81–0.87 for conversion from normal cognition to MCI. Longitudinal neuroimaging research has consistently demonstrated that structural brain atrophy precedes and closely tracks cognitive decline in AD [9, 18], with the OASIS dataset [29] serving as a widely adopted benchmark [12, 17].

### 2.3. Biomarker Trajectories and Predictive Performance

Lo et al. [20] analyzed 819 participants across 59 ADNI sites, examining trajectories of cerebrospinal fluid A*β*_42_, FDG-PET uptake, and hippocampal volume over 36 months. Glucose metabolic decline and hippocampal atrophy were significantly faster in AD than in normal participants (*p <* 0.001); hippocampal volume declined at a rate of −1.73%/year in AD versus −0.65%/year in cognitively normal individuals. Cognitive scores were best predicted by FDG changes in AD (partial *r*^2^ = 0.31), while both FDG and hippocampal changes contributed equally in MCI (partial *r*^2^ ≈ 0.18 each). Platero [15] used survival analysis with longitudinal data to predict MCI/dementia conversion, finding AUC = 0.82 for marker subsets combining temporal atrophy, clinical scores, and pTAU/A*β* ratios; relevant biomarker changes exceeding 20% were detectable approximately 15 years before cognitive decline onset, and Generalized Regression with Adaptive Component Estimation models outperformed longitudinal two-part joint mixed models with lower Akaike information criterion (AIC) values in cross-validated comparisons.

### 2.4. Machine Learning Approaches to Dementia Classification

A growing body of work has applied ML methods to dementia classification using neuroimaging and clinical data. Zhang et al. [23] proposed a multi-modal deep learning model integrating MRI features and clinical variables for AD diagnosis, reporting accuracy = 90.2%, sensitivity = 88.7%, and specificity = 91.4% on held-out test sets. Zhou et al. [24] developed a stage-wise deep neural network for multimodal dementia diagnosis that explicitly accounts for disease progression, achieving accuracy = 88.5% and AUC = 0.93 for AD versus MCI classification, demonstrating that temporal feature fusion improved F1-score by approximately 6 percentage points over cross-sectional baselines. Liu et al. [25] introduced a landmark-based deep multi-instance learning framework for brain lesion pattern classification, obtaining accuracy = 84.7% and AUC = 0.91 on the ADNI benchmark. Gradient-boosted ensemble classifiers [32] have achieved accuracy values in the range of 82–88% on tabular OASIS and ADNI features while offering superior interpretability over deep architectures. A comparison of reported performance across key studies is discussed in Section 5 and summarized in Table 24.

Sample sizes in the literature range from 120 [30] to 2,142 participants [8], with follow-up periods typically spanning 3–8 years, underscoring the diversity of study designs and the need for models robust to variable observation windows.

### 2.5. Clinical Assessment Instruments

The MMSE [10] and CDR [11] remain the most widely deployed instruments in dementia research and clinical practice. The CDR Sum of Boxes extension provides finer-grained staging sensitivity, particularly for early disease detection [33]. The MMSE, while sensitive to moderate-to-severe impairment, shows reduced discriminability at the MCI stage due to ceiling effects [10, 12]. Combined use of both instruments alongside structural imaging indicators has been recommended to improve early classification accuracy [5, 6].

### 2.6. Interpretability in Clinical Machine Learning

Interpretability has emerged as a critical requirement for Artificial Intelligence (AI)-assisted clinical tools [28]. SHAP [26], grounded in cooperative game theory, provides theoretically principled attribution scores that are consistent and locally accurate. Extensions including SHAP interaction values [27] enable the decomposition of model predictions into main effects and pairwise feature interactions, offering richer insight into the drivers of individual predictions. These properties make SHAP particularly well-suited for clinical dementia models, where understanding the contribution of specific biomarkers to a prediction is as important as the prediction itself [5, 28].

## 3. METHODS

### 3.1. Study Dataset

The OASIS-2 longitudinal MRI dataset [29], titled *Open Access Series of Imaging Studies (OASIS): Longitudinal MRI Data in Nondemented and Demented Older Adults*, was used as the primary data source for this study. The dataset is publicly available through the OASIS website https://www.oasis-brains.org (accessed March 16, 2026). The dataset was originally described by Marcus et al. [29] and contains longitudinal clinical and neuroimaging data for nondemented and demented older adults.

The dataset consisted of *N* = 150 unique subjects with repeated clinical and neuroimaging assessments, yielding a total of 373 observations (Table 1). Each subject had between 2 and 5 visits. At baseline, participants were aged between 60 and 96 years. Across all longitudinal visits included in the dataset, the recorded age range was 60 to 98 years.

**Table 1.** Summary of the OASIS-2 longitudinal dataset used in this study.

| Characteristic | Value |
| --- | --- |
| Total subjects | 150 |
| Total observations | 373 |
| Number of repeated visits per subject | 2–5 |
| Baseline age range | 60–96 years |
| Age range across all visits | 60–98 years |
| Non-demented observations | 190 |
| Demented observations | 146 |
| Converted observations | 37 |
| Outcome classes | Converted, Demented, Non-demented |

Let *i* = 1, … , *N* index subjects and *t* = 1, … , *T*_*i*_ index repeated visits for subject *i*. The observation for subject *i* at visit *t* is denoted by **x**_*it*_, and the corresponding diagnostic outcome is denoted by *y*_*it*_.

The diagnostic outcome consisted of three classes:

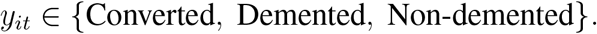

The final dataset contained 190 Non-demented observations, 146 Demented observations, and 37 Converted observations. Observations were sorted chronologically within each subject using the subject identifier, visit identifier, and time variable before longitudinal features were constructed.

### 3.2. Patient-Level Data Partitioning

To avoid data leakage across repeated observations, data partitioning was performed at the patient level rather than the observation level. This ensured that all visits belonging to a given patient appeared either in the training set or in the testing set, but not in both. A patient-level holdout split was used, with 80% of subjects assigned to training and 20% assigned to testing (Table 2). This resulted in 120 training patients and 30 testing patients, corresponding to 297 training observations and 76 testing observations.

**Table 2.** Patient-level training and testing split.

| Subset | Number of Subjects | Number of Observations |
| --- | --- | --- |
| Training set | 120 | 297 |
| Testing set | 30 | 76 |
| Total | 150 | 373 |

In addition to the held-out test evaluation, repeated stratified group cross-validation was used to assess internal performance stability. This cross-validation procedure preserved patient-level grouping so that repeated visits from the same subject did not appear in both training and validation folds.

### 3.3. Clinical and Neuroimaging Features

The current-visit feature set included seven variables: Age, MRI delay (`mr_delay`), Mini-Mental State Examination (MMSE) , Clinical Dementia Rating (CDR), estimated total intracranial volume (eTIV), normalized whole-brain volume (nWBV), and atlas scaling factor (ASF). The current-visit feature vector was defined as:

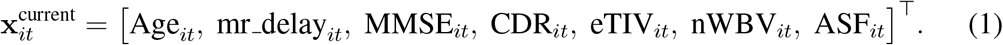

MMSE is a global cognitive screening measure [10], while CDR reflects clinical dementia severity [11]. The neuroimaging measures eTIV, nWBV, and ASF provide structural brain and head-size-related information. All the features are available in the OASIS-2 dataset [29].

### 3.4. Longitudinal Feature Engineering

To capture temporal disease progression, two additional types of longitudinal features were generated: previous-visit features and slope-based features. For each subject, previous-visit values were computed using the immediately preceding visit. For a longitudinal variable *z* ∈ *{*MMSE, CDR, nWBV, eTIV, ASF*}*, the previous-visit feature was defined as:

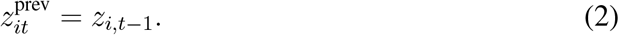

The rate of change between visits was computed using:

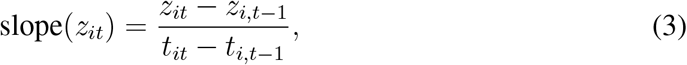

where *t*_*it*_ represents the time value for subject *i* at visit *t*. If the previous visit was unavailable or if the time difference was zero, the slope value was treated as missing. Missing values in all feature sets were handled using median imputation within the modeling pipeline.

Three feature sets were evaluated in this study, as presented in Table 3. The first used only current-visit features. The second added previous-visit features for MMSE, CDR, nWBV, eTIV, and ASF. The third used the full temporal feature set by adding slope-based features for the same five longitudinal variables. The final temporal feature set contained 17 predictors.

**Table 3.**
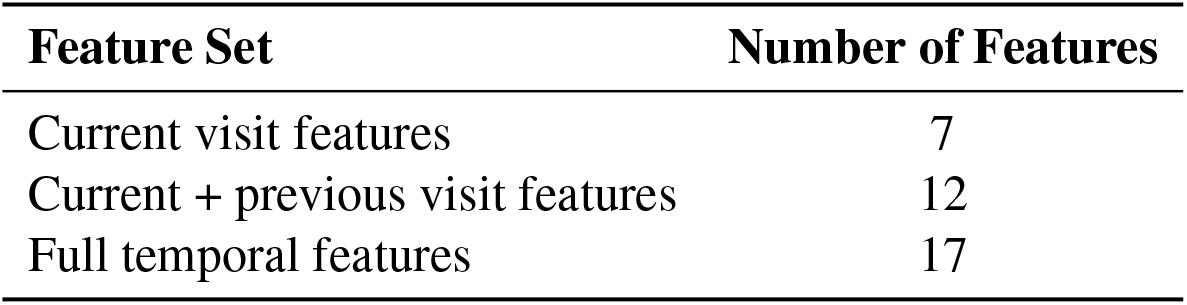
Feature sets evaluated in the study.

| Feature Set | Number of Features |
| --- | --- |
| Current visit features | 7 |
| Current + previous visit features | 12 |
| Full temporal features | 17 |

The current-visit feature set included Age, mr delay, MMSE, CDR, eTIV, nWBV, and ASF. The current plus previous-visit feature set added previous MMSE, CDR, nWBV, eTIV, and ASF. The full temporal feature set further included slope-based features for MMSE, CDR, nWBV, eTIV, and ASF.

### 3.5. Longitudinal Mixed-Effects Modeling

Before classification, linear mixed-effects models were fitted to examine longitudinal trajectories of cognitive decline and brain-volume change. Random intercept models were used to account for repeated measurements within subjects. Separate models were fitted for MMSE and nWBV.

The general mixed-effects model was specified as:

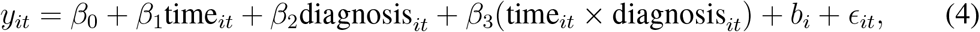

where *y*_*it*_ represents either MMSE or nWBV, *b*_*i*_ is the subject-specific random intercept, and *ϵ*_*it*_ is the residual error term. These models were used to assess whether cognitive and neuroimaging variables exhibited longitudinal patterns across diagnostic groups.

### 3.6. Baseline and Proposed Machine Learning Models

Several models were evaluated to determine whether longitudinal temporal feature engineering improved classification performance. The baseline models included a majority-class classifier, logistic regression, random foreGradientgradient boosting using current-visit features. The proposed model was a longitudinal gradient boosting classifier trained using the full temporal feature set.

The following models were evaluated:

1. Majority-class baseline using current-visit features;
2. Logistic regression baseline using current-visit features;
3. Random forest baseline using current-visit features;
4. Original longiGradientgradient boosting using full temporal features;
5. Regularized longiGradientgradient boosting using full temporal features.

The majority-class baseline was included to show the performance obtained by always predicting the dominant diagnostic class. Logistic regression was included as a linear baseline model. Random forest was included as a non-linear ensemble baseline. The longiGradientgradient boosting model was used as the proposed model because it can capture non-linear relationships and interactions among current, previous, and slope-based temporal predictors.

### 3.7. Gradient Boosting Model Specification

The proposed modelGradientgradient boosting classifier implemented using GradientBoostingClassifier from scikit-learn. Gradient boosting builds an additive ensemble of decision trees:

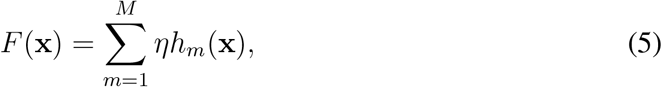

where *h*_*m*_(**x**) is the decision tree fitted at iteration *m, M* is the number of boosting estimators, and *η* is the learning rate.

For the three-class classification problem, the model optimized the multinomial deviance loss. The predicted probability for class *k* was computed using the softmax function:

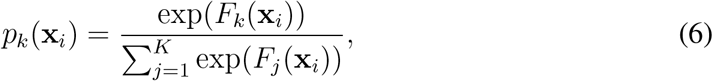

where *K* = 3 diagnostic classes.

Two longiGradientgradient boosting configurations were evaluated (Table 4). The oGra-dientgradient boosting model used 300 estimators, a learning rate of 0.05, a maximum tree depth of 3, and a subsampling rate of 0.80. A regularized version was also evaluated to examine overfitting. The regularized model used fewer estimators, a lower learning rate, shallower trees, larger minimum leaf size, larger minimum split size, and stronger subsampling.

**Table 4.** Hyperparameter settings for the longiGradientgradient boosting models.

| Model | n_estimators | learning_rate | max_depth | min_samples_leaf | min_samples_split | subsample |
| --- | --- | --- | --- | --- | --- | --- |
| Original longitudinal gradient boosting | 300 | 0.05 | 3 | Default | Default | 0.80 |
| Regularized longitudinal gradient boosting | 150 | 0.03 | 2 | 8 | 15 | 0.70 |

### 3.8. Handling Missing Values and Preprocessing

All models were implemented as scikit-learn pipelines (Table 5). Missing feature values, including undefined previous-visit and slope values for first visits, were handled using median imputation. Logistic regression additionally included standardization using `StandardScaler`. Tree-based models did not require feature scaling. The logistic regression model used a maximum of 1000 iterations and class balancing. The random forest model used 300 estimators, a maximum tree depth of 5, a minimum leaf size of 6, and balanced class weights. All models used a random seed of 42 where applicable.

**Table 5.** Implementation details for baseline and proposed models.

| Model | Implementation Details |
| --- | --- |
| Majority-class baseline | `DummyClassifier(strategy="most_frequent")` |
| Logistic regression | Median imputation, standardization, `max_iter=1000`, `class_weight="balanced"` |
| Random forest | 300 trees, `max_depth=5`, `min_samples_leaf=6`, `class_weight="balanced"` |
| OGradientgradient boosting | 300 estimators, learning rate 0.05, max depth 3, subsample 0.80 |
| ReguGradientgradient boosting | 150 estimators, learning rate 0.03, max depth 2, min leaf 8, min split 15, subsample 0.70 |

### 3.9. Model Evaluation Metrics

Model performance was evaluated on the held-out patient-level test set using accuracy, precision, recall, macro F1-score, and weighted F1-score. Accuracy was defined as:

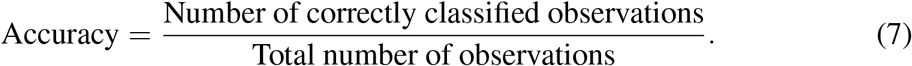

For each class *k*, precision, recall, and F1-score were computed as:

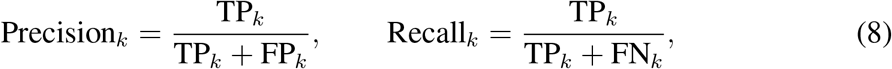

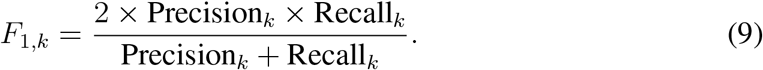

Macro F1-score was computed as the unweighted mean of class-specific F1-scores:

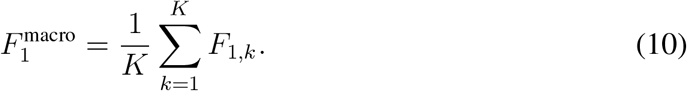

Weighted F1-score was computed by weighting each class-specific F1-score by its class support:

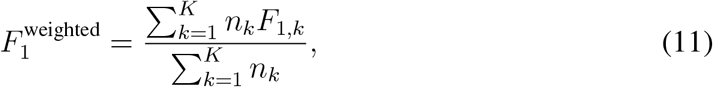

where *n*_*k*_ is the number of observations in class *k*.

Confusion matrices were generated for both the original and regularized longiGradient-gradient boosting models. The original model was treated as the main proposed model because it achieved the best held-out test performance, while the regularized model was treated as a sensitivity analysis for overfitting.

### 3.10. ROC AUC and Probability Calibration

Multiclass ROC curves were generated using a one-vs-rest approach. For each diagnostic class, the class was treated as the positive class and the remaining classes were treated as the negative class. The area under the ROC curve was computed for each class.

Calibration analysis was performed for the regularized longiGradientgradient boosting model using predicted class probabilities. One-vs-rest calibration curves were generated for each diagnostic class. Predicted probabilities were grouped into probability bins, and the observed class frequency was calculated within each bin. Bootstrap resampling with 1000 iterations was used to estimate 95% confidence intervals for observed calibration frequencies within bins.

Brier scores were computed for each class to quantify probability accuracy:

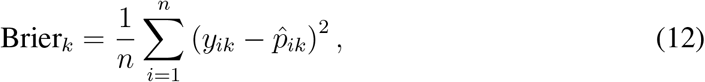

where *y*_*ik*_ is the binary indicator for class *k* and 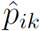 is the predicted probability for class *k*. Multiclass log loss was also computed to assess overall probability quality.

### 3.11. Bootstrap Confidence Intervals

Bootstrap confidence intervals were computed to quantify uncertainty in held-out test performance. The test set predictions were resampled with replacement 1000 times. For each bootstrap sample, accuracy, macro F1-score, and weighted F1-score were recalculated. The 2.5th and 97.5th percentiles of the bootstrap distribution were used as the lower and upper bounds of the 95% confidence interval.

### 3.12. Repeated Stratified Group Cross-Validation

To further assess internal model stability, repeated stratified group cross-validation was conducted using 5 folds repeated 5 times. Stratification preserved the diagnostic class distribution across folds, while grouping ensured that repeated visits from the same subject remained within the same fold. This approach was used to reduce leakage and provide a patient-level estimate of model stability across repeated train-test partitions.

For each fold, the regularized longiGradientgradient boosting model was trained on the training groups and evaluated on the held-out groups. Accuracy, macro F1-score, and weighted F1-score were recorded for each fold and summarized using the mean and standard deviation across the 25 folds.

### 3.13. Learning Curve and Overfitting Assessment

Learning curve analysis was performed to examine whether model performance improved with increasing training sample size and to assess overfitting. Group-based splitting was used so that repeated visits from the same patient were not split across training and validation subsets. Training sizes were evaluated at five levels, ranging from 20% to 100% of the available training data.

For each training size, macro F1-score was computed on the training and validation sets. The generalization gap was defined as:

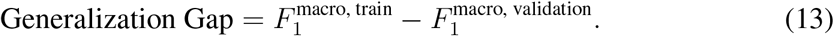

The original longiGradientgradient boosting model was compared with the regularized version to assess whether reducing model complexity reduced evidence of overfitting. The original model was retained as the main proposed model because it achieved the strongest held-out test performance, while the regularized model was reported as an overfitting sensitivity analysis.

### 3.14. Feature Importance and Permutation Importance

Model-based feature importance was extracted from the reguGradientgradient boosting classifier. This quantified the relative contribution of each predictor based on the total reduction in impurity attributed to splits involving that feature.

Permutation importance was also computed on the held-out test set using macro F1-score as the scoring metric. For each feature, values were randomly permuted multiple times, and the reduction in macro F1-score was recorded. A larger decrease in macro F1-score indicated that the feature contributed more strongly to classification performance.

### 3.15. SHAP-Based Interpretability

SHAP analysis was performed to provide class-specific model explanations. SHAP values were computed using a model-agnostic probability prediction function based on the final longiGradientgradient boosting model. A background sample from the training set was used to estimate baseline predictions, and SHAP values were generated for the held-out test observations.

For a prediction function *f* (**x**), the SHAP value for feature *j* is defined as:

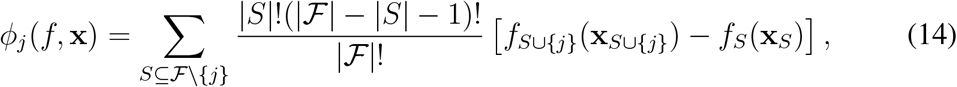

where *F* is the full set of features and *S* is a subset of features excluding feature *j*.

Class-specific SHAP summary plots and mean absolute SHAP bar plots were generated for the Converted, Demented, and Non-demented classes. These plots were used to identify features that increased or decreased the predicted probability of each diagnostic class.

### 3.16. Software and Reproducibility

All analyses were implemented in Python. The analysis used Python 3.13.9, scikit-learn 1.7.2, NumPy 2.3.5, and Pandas 2.3.3 (Table 6). The operating system was macOS-26.3 on ARM architecture. Linear mixed-effects models were fitted using the `statsmodels` package. Model interpretation used feature importance, permutation importance, and SHAP. A fixed random seed of 42 was used for data partitioning, model fitting where applicable, bootstrap resampling, and cross-validation procedures.

**Table 6.** Software and reproducibility information.

| Item | Value |
| --- | --- |
| Python version | 3.13.9 |
| scikit-learn version | 1.7.2 |
| NumPy version | 2.3.5 |
| Pandas version | 2.3.3 |
| Operating system | macOS-26.3-arm64 |
| Processor architecture | ARM |
| Random seed | 42 |

## 4. RESULTS

### 4.1. Reproducibility and Dataset Description

All analyses were conducted using Python 3.13.9 with scikit-learn 1.7.2, NumPy 2.3.5, and Pandas 2.3.3. A fixed random seed of 42 was used to support reproducibility. The dataset contained 373 longitudinal observations from 150 patients. As shown in Table 8, the diagnostic classes were Non-demented, Demented, and Converted. The largest class was Non-demented with 190 observations, followed by Demented with 146 observations and Converted with 37 observations.

Three feature sets were evaluated. The current-visit feature set contained 7 predictors: Age, mr delay, MMSE, CDR, eTIV, nWBV, and ASF. The current plus previous-visit feature set contained 12 predictors after adding previous values of MMSE, CDR, nWBV, eTIV, and ASF. The full temporal feature set contained 17 predictors after further adding slope-based features for MMSE, CDR, nWBV, eTIV, and ASF. These slope variables represented the rate of change between the current and previous patient visit.

To avoid leakage across repeated visits, the data were split at the patient level. A total of 120 patients were assigned to the training set and 30 patients to the testing set. This corresponded to 297 training observations and 76 testing observations, as summarized in Table 7.

**Table 7.** Summary of dataset characteristics.

| Characteristic | Value |
| --- | --- |
| Total observations | 373 |
| Total patients | 150 |
| Training patients | 120 |
| Testing patients | 30 |
| Training observations | 297 |
| Testing observations | 76 |
| Current-visit features | 7 |
| Current + previous-visit features | 12 |
| Full temporal features | 17 |

**Table 8.** Diagnostic class distribution.

| Diagnosis | Number of Observations |
| --- | --- |
| Non-demented | 190 |
| Demented | 146 |
| Converted | 37 |

### 4.2. Longitudinal Mixed-Effects Analysis

Linear mixed-effects models were fitted to evaluate longitudinal trajectories in MMSE and nWBV while accounting for repeated observations within patients. A random intercept was included for each patient. The MMSE mixed-effects model is presented in Table 9. The model included 371 observations from 150 patient groups and converged successfully. The Demented group had significantly lower MMSE scores than the reference group, with an estimated coefficient of −3.798 (*p <* 0.001). This indicates that Demented participants had poorer cognitive performance. The coefficient for time was negative (*β* = −0.224), suggesting a decrease in MMSE over time, although this effect was not statistically significant (*p* = −0.125). The time-by-diagnosis interaction terms for Demented and Non-demented participants were also not statistically significant.

**Table 9.** Linear mixed-effects model for MMSE trajectory.

| Term | Coefficient | Std. Error | z-value | p-value | 95% CI |
| --- | --- | --- | --- | --- | --- |
| Intercept | 29.035 | 0.730 | 39.759 | $< 0.001$ | [27.604, 30.467] |
| Demented | -3.798 | 0.807 | -4.706 | $< 0.001$ | [-5.380, -2.216] |
| Non-demented | 0.113 | 0.798 | 0.141 | 0.888 | [-1.451, 1.676] |
| Time | -0.224 | 0.146 | -1.535 | 0.125 | [-0.509, 0.062] |
| Time × Demented | -0.276 | 0.183 | -1.511 | 0.131 | [-0.634, 0.082] |
| Time × Non-demented | 0.248 | 0.164 | 1.515 | 0.130 | [-0.073, 0.569] |
| Group variance | 5.073 | 0.543 | — | — | — |

The nWBV mixed-effects model is presented in Table 10. The model included 373 observations from 150 patient groups and also converged successfully. Time had a significant negative effect on nWBV (*β* = −0.006, *p <* 0.001), indicating a measurable decline in normalized whole-brain volume over the follow-up period. The main effect for Demented status was negative but not statistically significant (*β* = −0.015, *p* = 0.144). The interaction between time and Non-demented status was positive and statistically significant (*β* = 0.002, *p* = 0.001), indicating that the rate of nWBV change differed across diagnostic groups.

**Table 10.** Linear mixed-effects model for nWBV trajectory.

| Term | Coefficient | Std. Error | z-value | p-value | 95% CI |
| --- | --- | --- | --- | --- | --- |
| Intercept | 0.739 | 0.009 | 78.361 | $< 0.001$ | [0.720, 0.757] |
| Demented | -0.015 | 0.010 | -1.462 | 0.144 | [-0.036, 0.005] |
| Non-demented | 0.007 | 0.010 | 0.702 | 0.483 | [-0.013, 0.027] |
| Time | -0.006 | 0.001 | -8.407 | $< 0.001$ | [-0.007, -0.004] |
| Time × Demented | 0.001 | 0.001 | 0.913 | 0.361 | [-0.001, 0.002] |
| Time × Non-demented | 0.002 | 0.001 | 3.219 | 0.001 | [0.001, 0.004] |
| Group variance | 0.001 | 0.023 | — | — | — |

The observed trajectory plots supported the use of longitudinal analysis. Figure 1 shows the observed cognitive trajectory over time using MMSE, while Figure 2 shows the observed change in normalized whole-brain volume over time.

**Figure 1.**
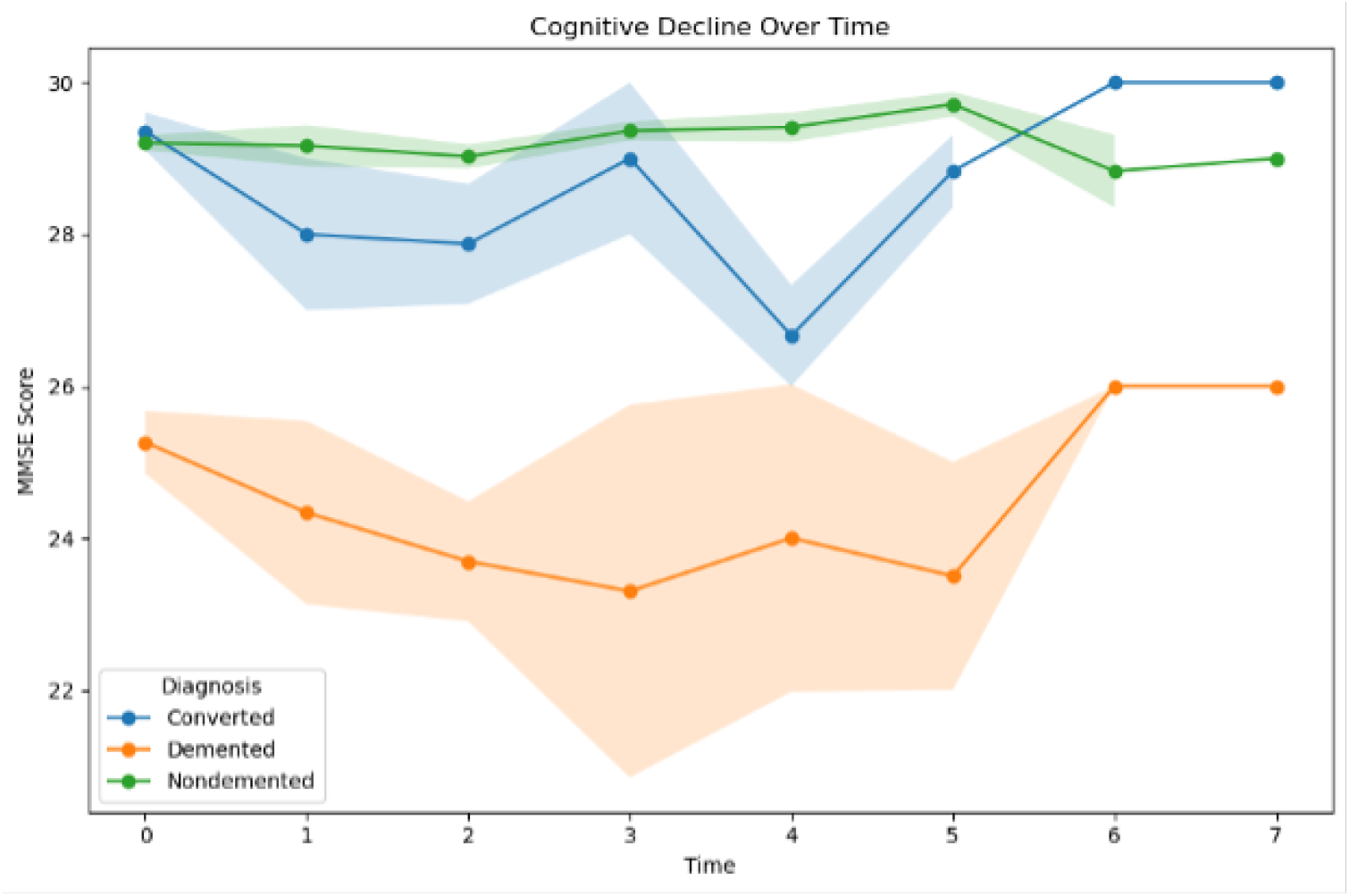
Observed cognitive decline over time based on MMSE. The plot shows mean MMSE values across time by diagnostic group, with variability represented by error bands.

**Figure 2.**
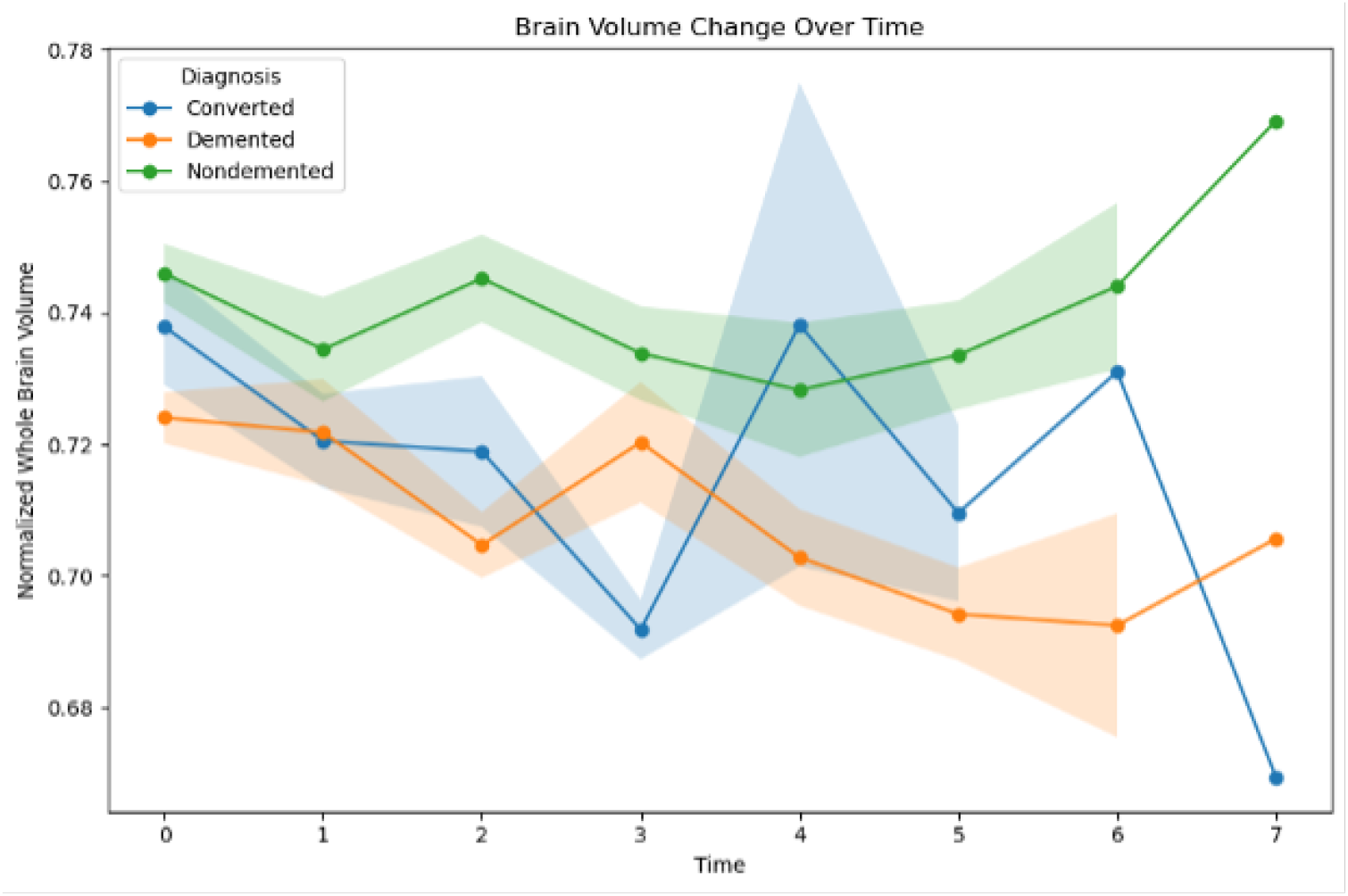
Observed brain volume change over time based on normalized whole-brain volume. The plot shows mean nWBV values across time by diagnostic group, with variability represented by error bands.

### 4.3. Model Performance and Overfitting Assessment

Five models were evaluated: a majority-class baseline, logistic regression, random forest, original longiGradientgradient boosting, and regularized longiGradientgradient boosting. Their training and test performance are summarized in Table 11. The majority-class baseline performed poorly, with a test accuracy of 0.6184 and a test macro F1-score of 0.2547. Logistic regression improved class-balanced performance, achieving a test accuracy of 0.7632 and a test macro F1-score of 0.7101. Random forest achieved a test accuracy of 0.8026 but had a lower macro F1-score of 0.6160, suggesting weaker performance across all classes.

**Table 11.** Model performance and overfitting assessment.

| Model | Features | Train Acc. | Test Acc. | Train Macro F1 | Test Macro F1 | Train Weighted F1 | Test Weighted F1 | Overfitting Gap |
| --- | --- | --- | --- | --- | --- | --- | --- | --- |
| Majority class baseline | 7 | 0.4815 | 0.6184 | 0.2167 | 0.2547 | 0.3130 | 0.4726 | -0.0381 |
| Logistic regression baseline | 7 | 0.8586 | 0.7632 | 0.7479 | 0.7101 | 0.8747 | 0.7760 | 0.0378 |
| Random forest baseline | 7 | 0.9529 | 0.8026 | 0.8898 | 0.6160 | 0.9516 | 0.7620 | 0.2737 |
| Original longitudinal gradient boosting | 17 | 1.0000 | 0.8816 | 1.0000 | 0.7761 | 1.0000 | 0.8604 | 0.2239 |
| Regularized longitudinal gradient boosting | 17 | 0.9697 | 0.8421 | 0.9163 | 0.6815 | 0.9667 | 0.8060 | 0.2348 |

The original longiGradientgradient boosting model achieved the highest held-out test performance. It produced a test accuracy of 0.8816, a test macro F1-score of 0.7761, and a test weighted F1-score of 0.8604. However, its training accuracy, training macro F1-score, and training weighted F1-score were all 1.0000, indicating a perfect fit on the training data and suggesting possible overfitting.

To address this, a regularized gradient boosting model was fitted with fewer estimators, a lower learning rate, a smaller tree depth, a larger minimum leaf size, a larger minimum split size, and stronger subsampling. The regularized model reduced the training macro F1-score from 1.0000 to 0.9163, indicating that it no longer fit the training data perfectly. However, the test performance decreased to 0.8421 accuracy, 0.6815 macro F1-score, and 0.8060 weighted F1-score. Therefore, regularization reduced the perfect training fit but did not improve held-out test performance in this cohort.

### 4.4. Class-Specific Classification Results

The original longiGradientgradient boosting model performed best overall and showed strong classification performance for the Demented and Non-demented classes, as shown in Table 12. For the Demented class, the model achieved a precision of 0.85, a recall of 1.00, and an F1-score of 0.92. For the Non-demented class, precision was 0.90, recall was 0.98, and F1-score was 0.94. The Converted class was more difficult to classify, with a precision of 0.80, a recall of 0.33, and an F1-score of 0.47.

**Table 12.** Class-specific performance of the original longiGradientgradient boosting model.

| Class | Precision | Recall | F1-score | Support |
| --- | --- | --- | --- | --- |
| Converted | 0.80 | 0.33 | 0.47 | 12 |
| Demented | 0.85 | 1.00 | 0.92 | 17 |
| Non-demented | 0.90 | 0.98 | 0.94 | 47 |
| Accuracy |  | 0.8816 |  | 76 |
| Macro average | 0.85 | 0.77 | 0.78 | 76 |
| Weighted average | 0.87 | 0.88 | 0.86 | 76 |

The regularized model preserved perfect recall for the Demented class but reduced performance for the Converted class, as shown in Table 13. Converted recall decreased from 0.33 in the original model to 0.17 in the regularized model, and Converted F1-score decreased from 0.47 to 0.27. This indicates that regularization did not improve the classification of the transitional Converted group.

**Table 13.**
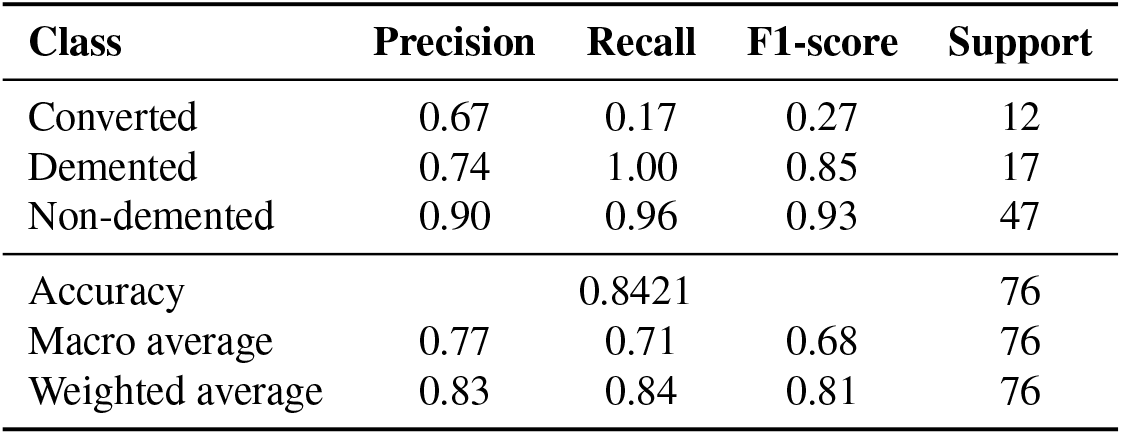
Class-specific performance of the regularized longiGradientgradient boosting model.

### 4.5. Confusion Matrix Results

The confusion matrix for the original longiGradientgradient boosting model is shown in Table 14 and Figure 3. The model correctly classified 4 of the 12 Converted cases. Three Converted cases were classified as Demented, and five were classified as Non-demented. All 17 Demented cases were correctly classified. For the Non-demented class, 46 of 47 cases were correctly classified, with one case misclassified as Converted.

**Table 14.**
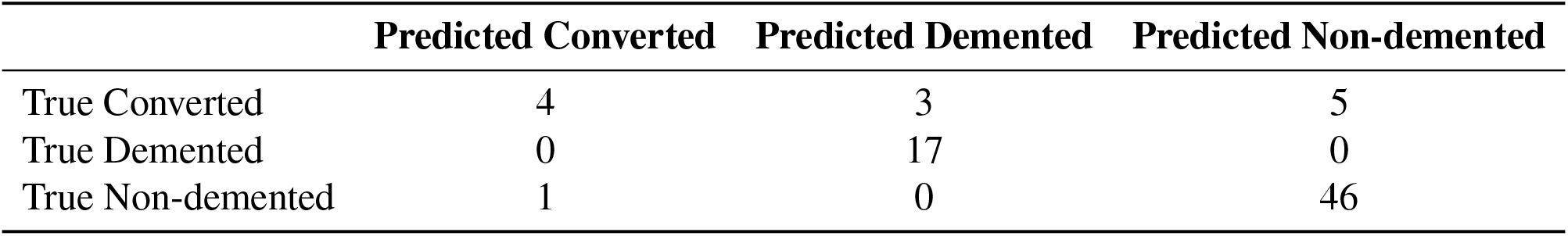
Confusion matrix for the original longiGradientgradient boosting model.

|  | Predicted Converted | Predicted Demented | Predicted Non-demented |
| --- | --- | --- | --- |
| True Converted | 4 | 3 | 5 |
| True Demented | 0 | 17 | 0 |
| True Non-demented | 1 | 0 | 46 |

**Figure 3.**
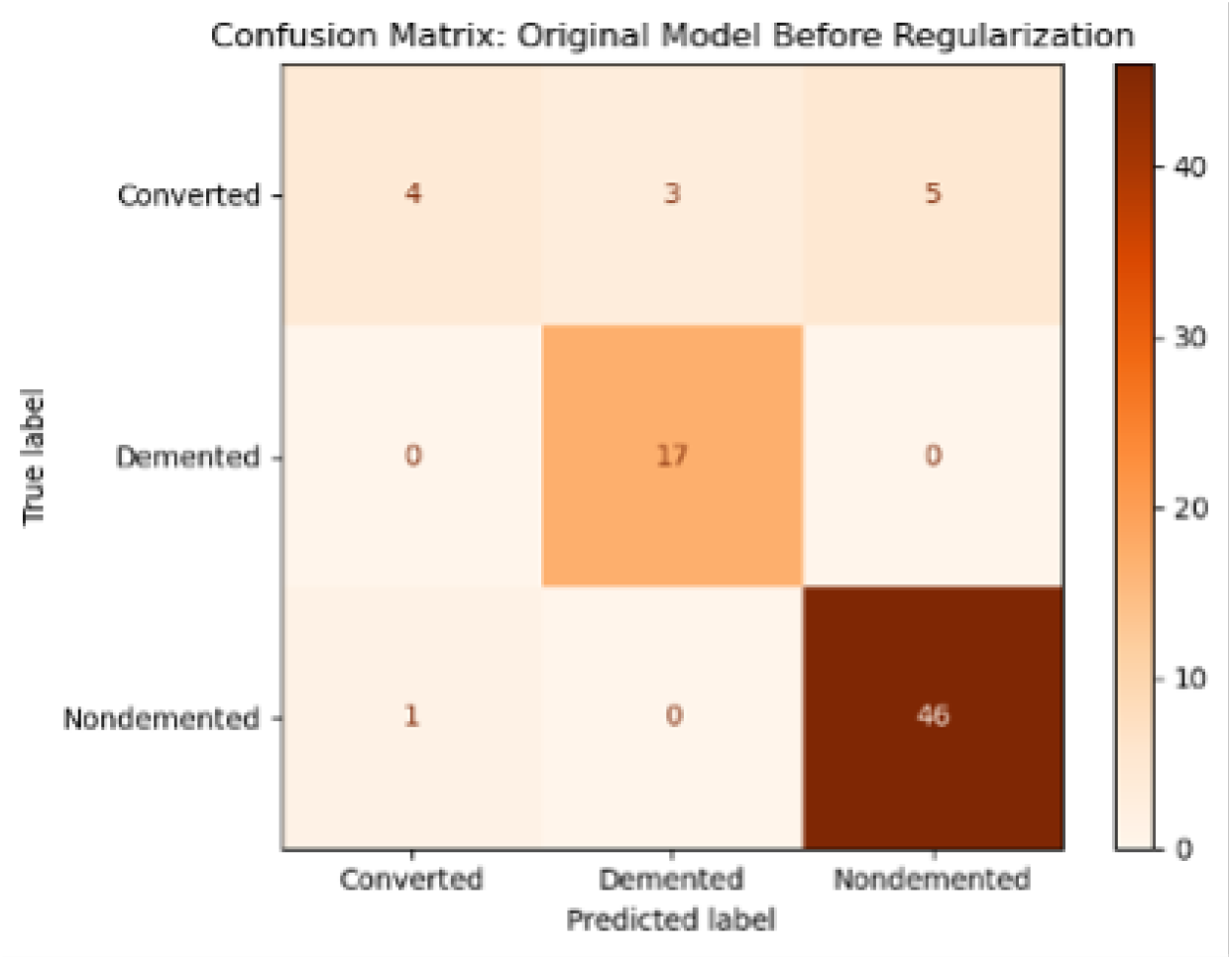
Confusion matrix for the original longiGradientgradient boosting model before regularization.

The confusion matrix for the regularized longiGradientgradient boosting model is shown in Table 15 and Figure 4. The regularized model correctly classified 2 of 12 Converted cases. Five Converted cases were classified as Demented, and five were classified as Non-demented. All 17 Demented cases were correctly classified again. For the Non-demented class, 45 of 47 cases were correctly classified, with one case classified as Converted and one as Demented.

**Table 15.**
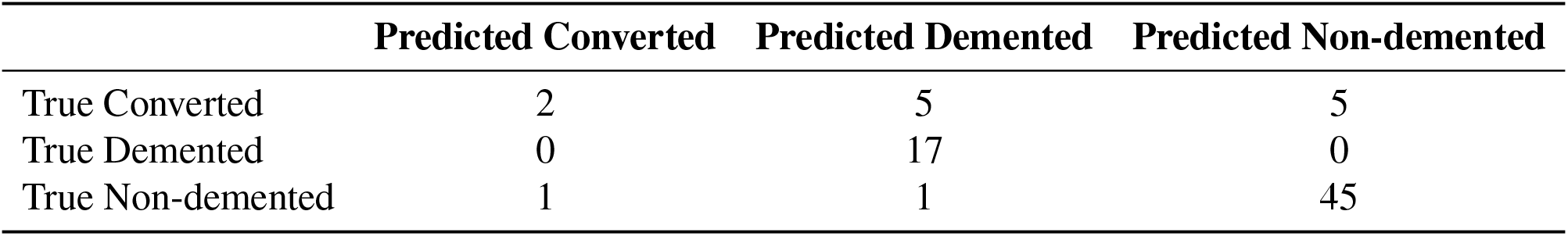
Confusion matrix for the regularized longitudinal Gradient boosting model.

**Figure 4.**
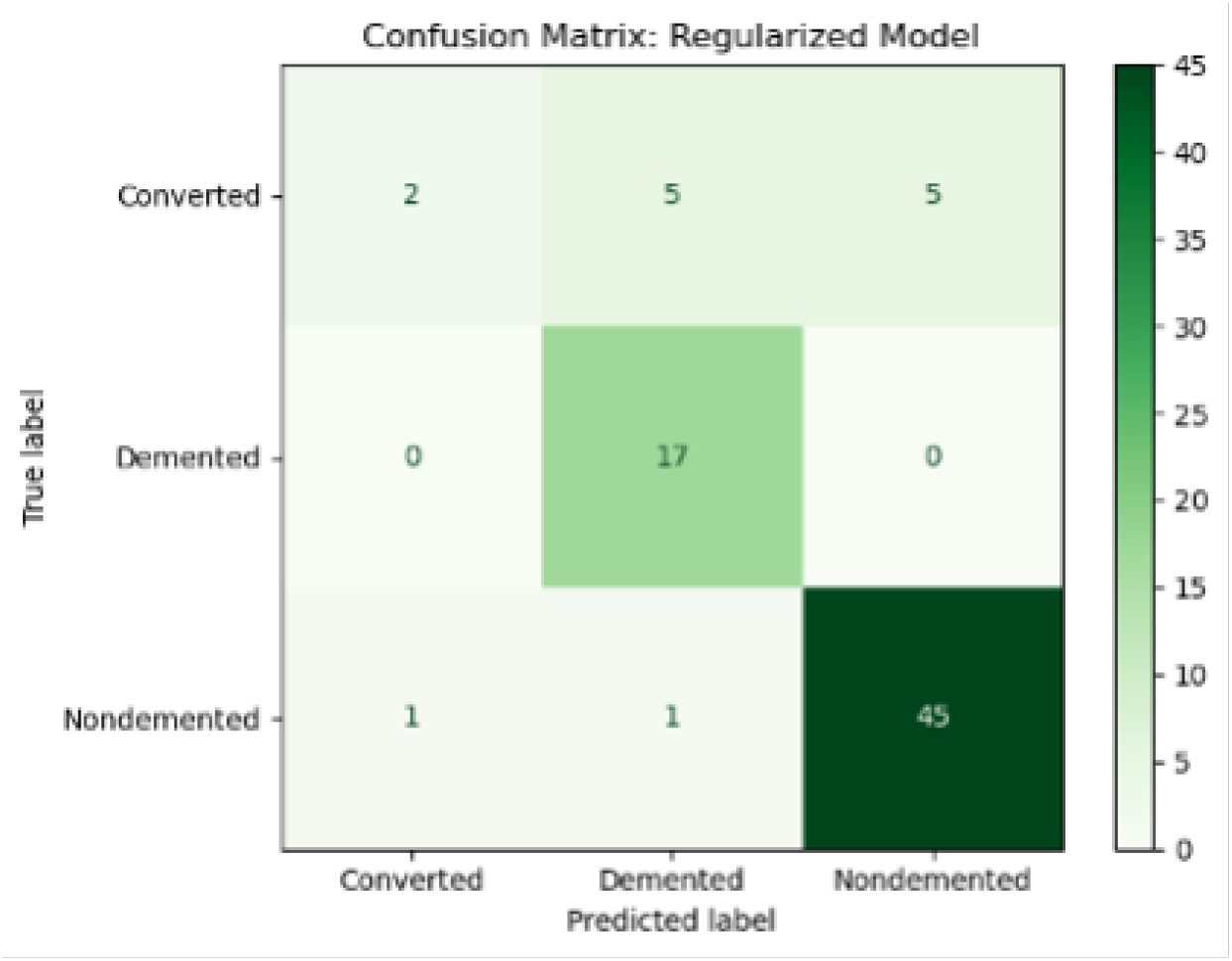
Confusion matrix for the regularized longitudinal Gradient boosting model.

These confusion matrices show that the main source of classification error was the Converted class. This class represents a transitional diagnostic group and was less frequent in the test set than the Non-demented class.

### 4.6. Bootstrap Confidence Intervals

Bootstrap confidence intervals were calculated for the regularized longitudinal Gradient boosting model, as shown in Table 16. The bootstrap mean accuracy was 0.8404, with a 95% confidence interval of 0.7632 to 0.9211. The bootstrap mean macro F1-score was 0.6757, with a 95% confidence interval of 0.5658 to 0.8062. The bootstrap mean weighted F1-score was 0.8034, with a 95% confidence interval of 0.7009 to 0.9030.

**Table 16.** Bootstrap 95% confidence intervals for the regularized longitudinal Gradient boosting model.

| Metric | Mean | Lower 95% CI | Upper 95% CI |
| --- | --- | --- | --- |
| Accuracy | 0.8404 | 0.7632 | 0.9211 |
| Macro F1-score | 0.6757 | 0.5658 | 0.8062 |
| Weighted F1-score | 0.8034 | 0.7009 | 0.9030 |

The macro F1-score had a wider interval than accuracy, reflecting greater uncertainty in class-balanced performance. This is consistent with the smaller number of Converted observations and the weaker performance for this class.

### 4.7. Repeated Stratified Group Cross-Validation

Repeated stratified group cross-validation was performed using 5 folds repeated 5 times. This approach preserved patient-level grouping so that repeated visits from the same patient were not split across training and validation folds. The cross-validation summary is shown in Table 17. Across the 25 validation folds, the regularized longitudinal Gradient boosting model achieved a mean accuracy of 0.9211 with a standard deviation of 0.0359. The mean macro F1-score was 0.7755 with a standard deviation of 0.0555, while the mean weighted F1-score was 0.9040 with a standard deviation of 0.0462.

**Table 17.** Repeated stratified group cross-validation summary for the regularized model.

| Metric | Mean | Standard Deviation |
| --- | --- | --- |
| Accuracy | 0.9211 | 0.0359 |
| Macro F1-score | 0.7755 | 0.0555 |
| Weighted F1-score | 0.9040 | 0.0462 |

These cross-validation results suggest that the regularized model exhibited stable internal validation performance under patient-level grouping, although the held-out test set performance remained below the repeated cross-validation mean.

### 4.8. ROC AUC Analysis

A multiclass ROC analysis was performed using a one-vs-rest approach for the regularized model. The class-specific AUC values are shown in Table 18, and the ROC curves are shown in Figure 5. The Demented class achieved the highest AUC of 0.9950, indicating strong discrimination between Demented and non-Demented cases. The Non-demented class also showed good discrimination, with an AUC of 0.8804. The Converted class had the lowest AUC of 0.6289, indicating weaker separation of transitional cases from the other diagnostic groups.

**Table 18.** Class-specific ROC AUC values for the regularized longitudinal Gradient boosting model.

| Class | AUC |
| --- | --- |
| Converted | 0.6289 |
| Demented | 0.9950 |
| Non-demented | 0.8804 |

**Figure 5.**
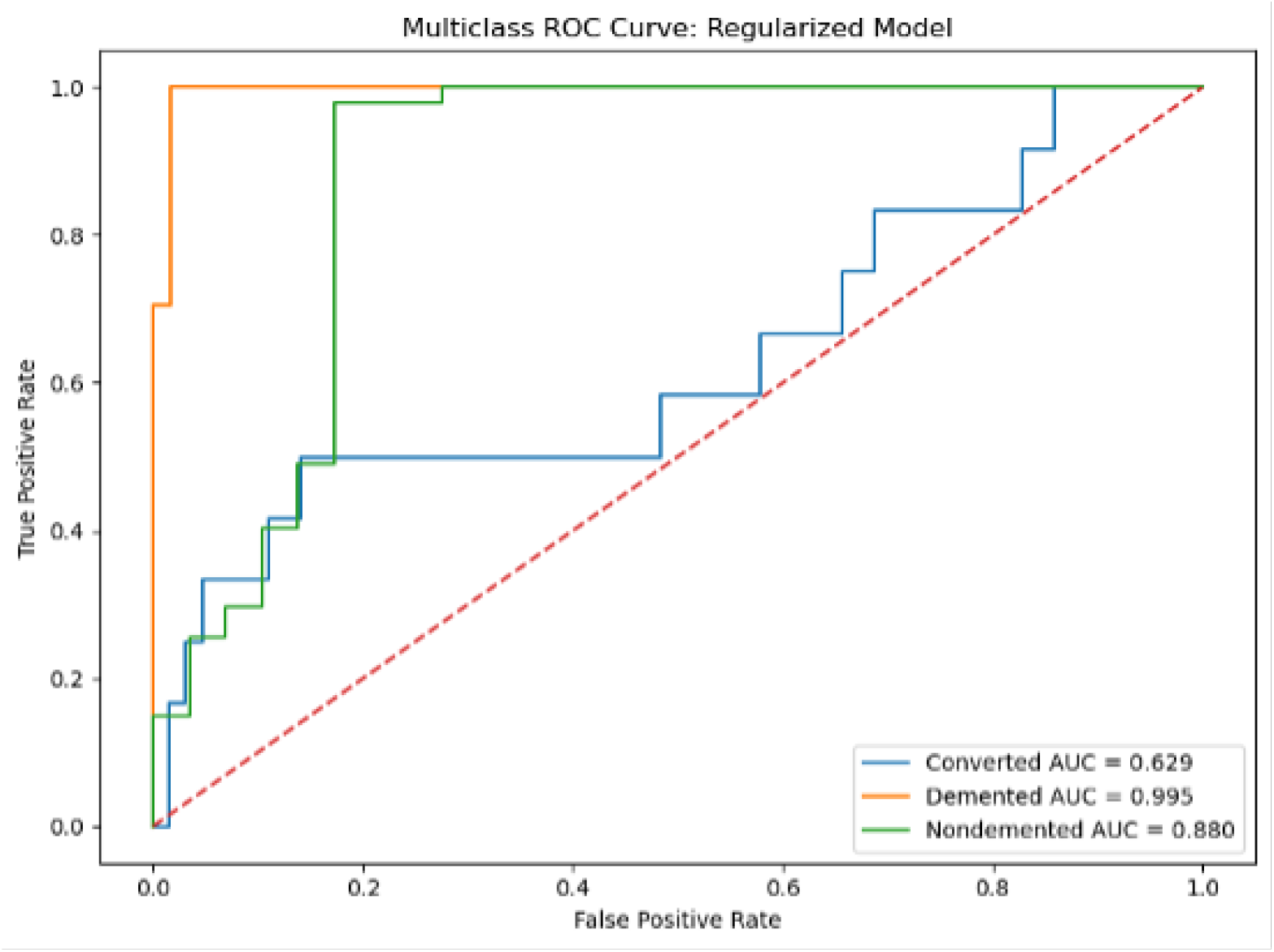
Multiclass ROC curve for the regularized longitudinal Gradient boosting model using a one-vs-rest approach.

### 4.9. Calibration Analysis

Calibration analysis was performed for the regularized model using predicted class probabilities. The calibration metrics are summarized in Table 19. The Brier score was lowest for the Demented class, with a value of 0.0449. The Non-demented class had a Brier score of 0.0885, while the Converted class had the highest Brier score of 0.1285. The multiclass log loss was 0.5355. These findings suggest that predicted probabilities were more reliable for the Demented and Non-demented classes than for the Converted class.

**Table 19.** Probability calibration metrics for the regularized longitudinal Gradient boosting model.

| Class/Metric | Value |
| --- | --- |
| Converted Brier score | 0.1285 |
| Demented Brier score | 0.0449 |
| Non-demented Brier score | 0.0885 |
| Multiclass log loss | 0.5355 |

Calibration points with bootstrap 95% confidence intervals are shown in Table 20. Wider confidence intervals were observed in bins with small sample counts, reflecting higher uncertainty in observed class frequencies. This was particularly important for the Converted class because of its smaller number of test observations and weaker classification performance.

**Table 20.** Calibration points with 95% confidence intervals for the regularized model.

| Class | Bin | Mean Predicted Probability | Observed Fraction | 95% CI | Bin Count |
| --- | --- | --- | --- | --- | --- |
| Converted | 1 | 0.0414 | 0.1176 | [0.0441, 0.1912] | 68 |
| Converted | 2 | 0.3742 | 0.3333 | [0.0000, 1.0000] | 3 |
| Converted | 3 | 0.4404 | 0.7500 | [0.2500, 1.0000] | 4 |
| Converted | 4 | 0.7168 | 0.0000 | [0.0000, 0.0000] | 1 |
| Demented | 1 | 0.0159 | 0.0000 | [0.0000, 0.0000] | 51 |
| Demented | 3 | 0.4875 | 0.0000 | [0.0000, 0.0000] | 5 |
| Demented | 4 | 0.7379 | 0.0000 | [0.0000, 0.0000] | 1 |
| Demented | 5 | 0.9649 | 0.8947 | [0.7368, 1.0000] | 19 |
| Non-demented | 1 | 0.0327 | 0.0400 | [0.0000, 0.1200] | 25 |
| Non-demented | 2 | 0.2653 | 1.0000 | [1.0000, 1.0000] | 1 |
| Non-demented | 3 | 0.5682 | 1.0000 | [1.0000, 1.0000] | 2 |
| Non-demented | 4 | 0.7940 | 1.0000 | [1.0000, 1.0000] | 2 |
| Non-demented | 5 | 0.9448 | 0.8913 | [0.8043, 0.9783] | 46 |

### 4.10. Feature Importance

Feature importance from the regularized Gradient boosting model is shown in Table 21 and Figure 6. CDR was the most influential predictor, with an importance value of 0.7789. This was much higher than all other predictors, indicating that current dementia severity was the dominant driver of model classification. The next most important variables were mr delay, Age, slope CDR, eTIV, ASF, and prev CDR. The presence of slope CDR among the top predictors suggests that temporal change in dementia severity contributed to classification beyond current CDR alone.

**Table 21.** Gradient boosting feature importance for the regularized model.

| Feature | Importance |
| --- | --- |
| CDR | 0.7789 |
| mr_delay | 0.0481 |
| Age | 0.0398 |
| slope_CDR | 0.0395 |
| eTIV | 0.0218 |
| ASF | 0.0174 |
| prev_CDR | 0.0155 |
| nWBV | 0.0101 |
| MMSE | 0.0095 |
| prev_MMSE | 0.0065 |
| slope_nWBV | 0.0047 |
| slope_MMSE | 0.0037 |
| prev_eTIV | 0.0013 |
| slope_eTIV | 0.0012 |
| prev_nWBV | 0.0011 |
| slope_ASF | 0.0005 |
| prev_ASF | 0.0004 |

**Figure 6.**
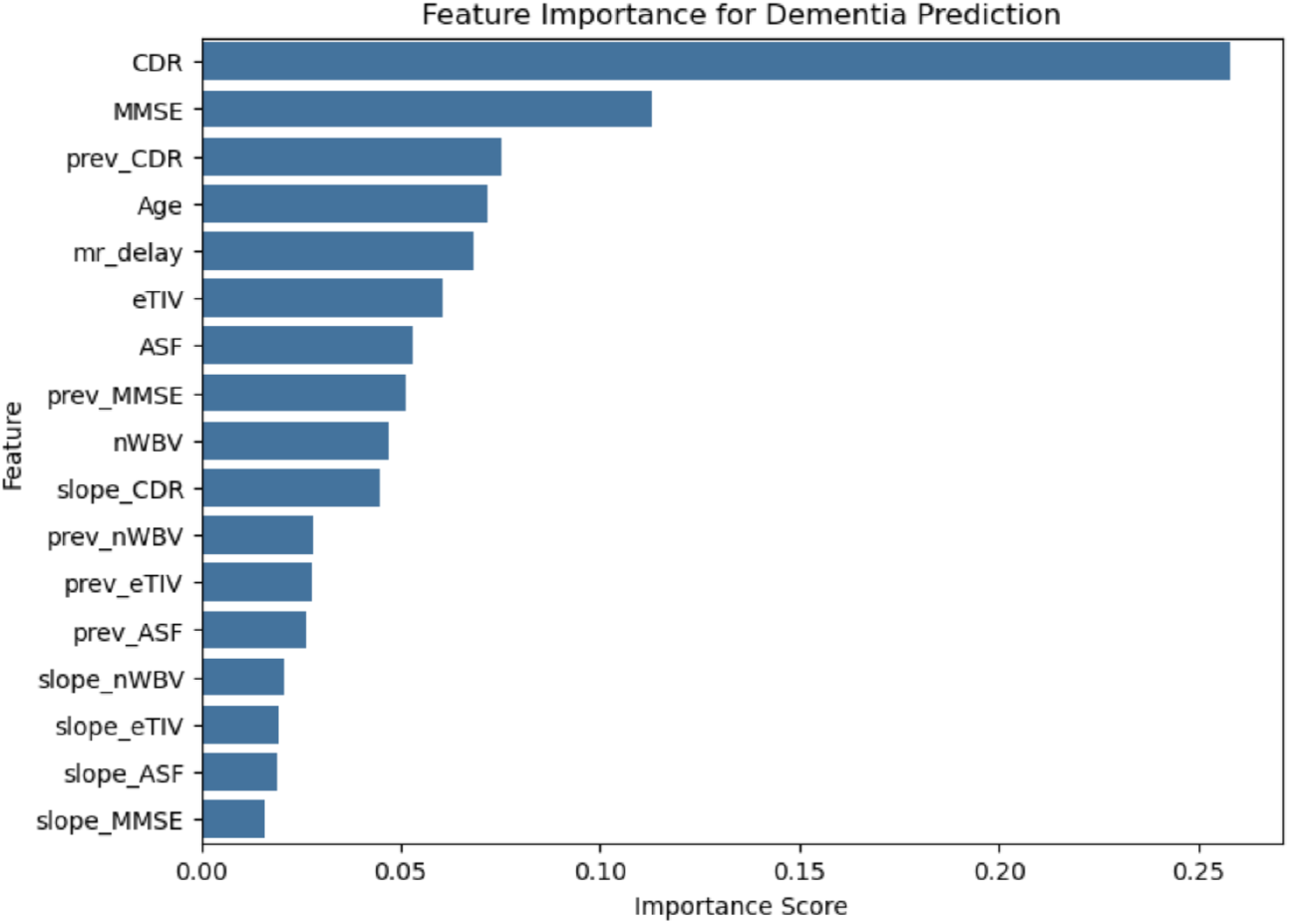
Top feature importance values from the regularized longitudinal Gradient boosting model

### 4.11. Permutation Importance

Permutation importance based on macro F1-score is summarized in Table 22 and Figure 7. Permuting CDR caused the largest decrease in macro F1-score, with a mean importance of 0.3616. The next most important feature was slope CDR, with a mean importance of 0.0888. Other features with positive permutation importance included mr delay, slope ASF, MMSE, nWBV, prev CDR, and prev MMSE.

**Table 22.** Permutation importance based on macro F1-score for the regularized model.

| Feature | Mean Importance | Standard Deviation |
| --- | --- | --- |
| CDR | 0.3616 | 0.0422 |
| slope_CDR | 0.0888 | 0.0254 |
| mr_delay | 0.0476 | 0.0380 |
| slope_ASF | 0.0369 | 0.0204 |
| MMSE | 0.0313 | 0.0349 |
| nWBV | 0.0289 | 0.0427 |
| prev_CDR | 0.0163 | 0.0265 |
| prev_MMSE | 0.0066 | 0.0202 |
| prev_eTIV | 0.0000 | 0.0000 |
| prev_ASF | 0.0000 | 0.0000 |
| slope_eTIV | 0.0000 | 0.0000 |
| slope_nWBV | -0.0013 | 0.0142 |
| prev_nWBV | -0.0043 | 0.0130 |
| slope_MMSE | -0.0048 | 0.0227 |
| Age | -0.0191 | 0.0397 |
| eTIV | -0.0385 | 0.0359 |
| ASF | -0.0472 | 0.0367 |

**Figure 7.**
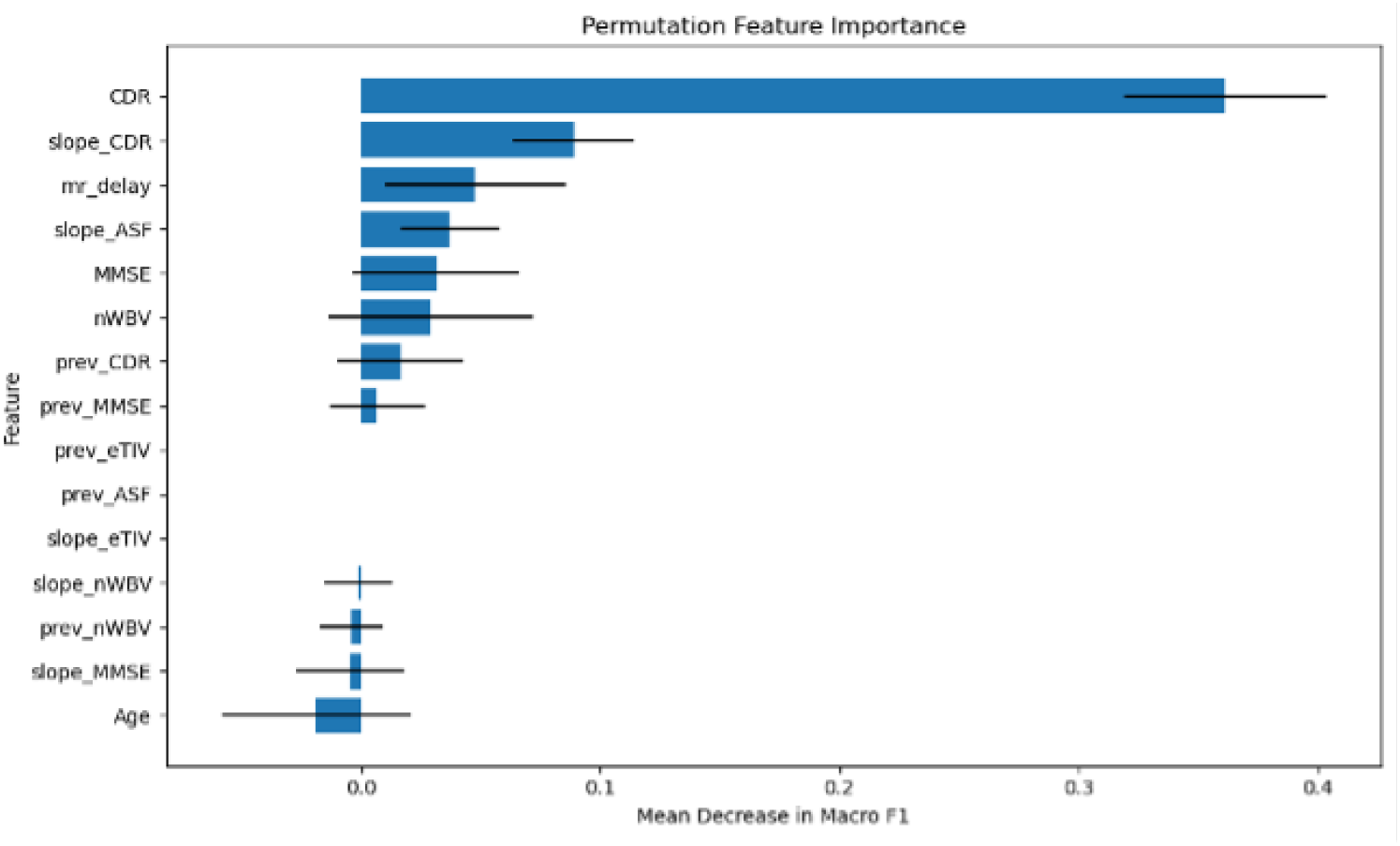
Permutation feature importance based on macro F1-score for the regularized longitudinal gradient boosting model.

Some features had near-zero or negative permutation importance, including eTIV, ASF, Age, slope MMSE, prev nWBV, and slope nWBV. Negative permutation importance suggests that permuting those variables did not reduce macro F1-score in the held-out test set and may reflect instability due to small sample size, correlation among predictors, or limited independent contribution after accounting for stronger features such as CDR.

### 4.12. SHAP-Based Model Explainability and Clinical Interpretation

SHAP analysis was used to interpret the final longitudinal Gradient boosting model. Class-specific SHAP summary plots and mean absolute SHAP plots were generated for the Converted, Demented, and Non-demented classes. The SHAP summary plots show the direction and spread of each feature’s effect on model prediction, while the mean absolute SHAP plots rank features according to their average contribution to each class prediction. Positive SHAP values indicate that a feature increased the predicted probability of a class, whereas negative SHAP values indicate that a feature decreased it.

For the Converted class, slope CDR had the largest mean absolute SHAP value, approximately 0.05, followed by Age at approximately 0.04. CDR, eTIV, mr delay, and ASF each contributed approximately 0.02, while MMSE, nWBV, and slope MMSE had smaller contributions of approximately 0.01. The SHAP summary plot in Figure 8 showed that higher slope CDR values tended to push predictions toward the Converted class, while lower slope CDR values generally reduced the probability of Converted classification. The mean absolute SHAP plot in Figure 9 confirmed that slope CDR was the most important feature for this class. This suggests that change in dementia severity over time was the most informative feature for identifying transitional cases.

**Figure 8.**
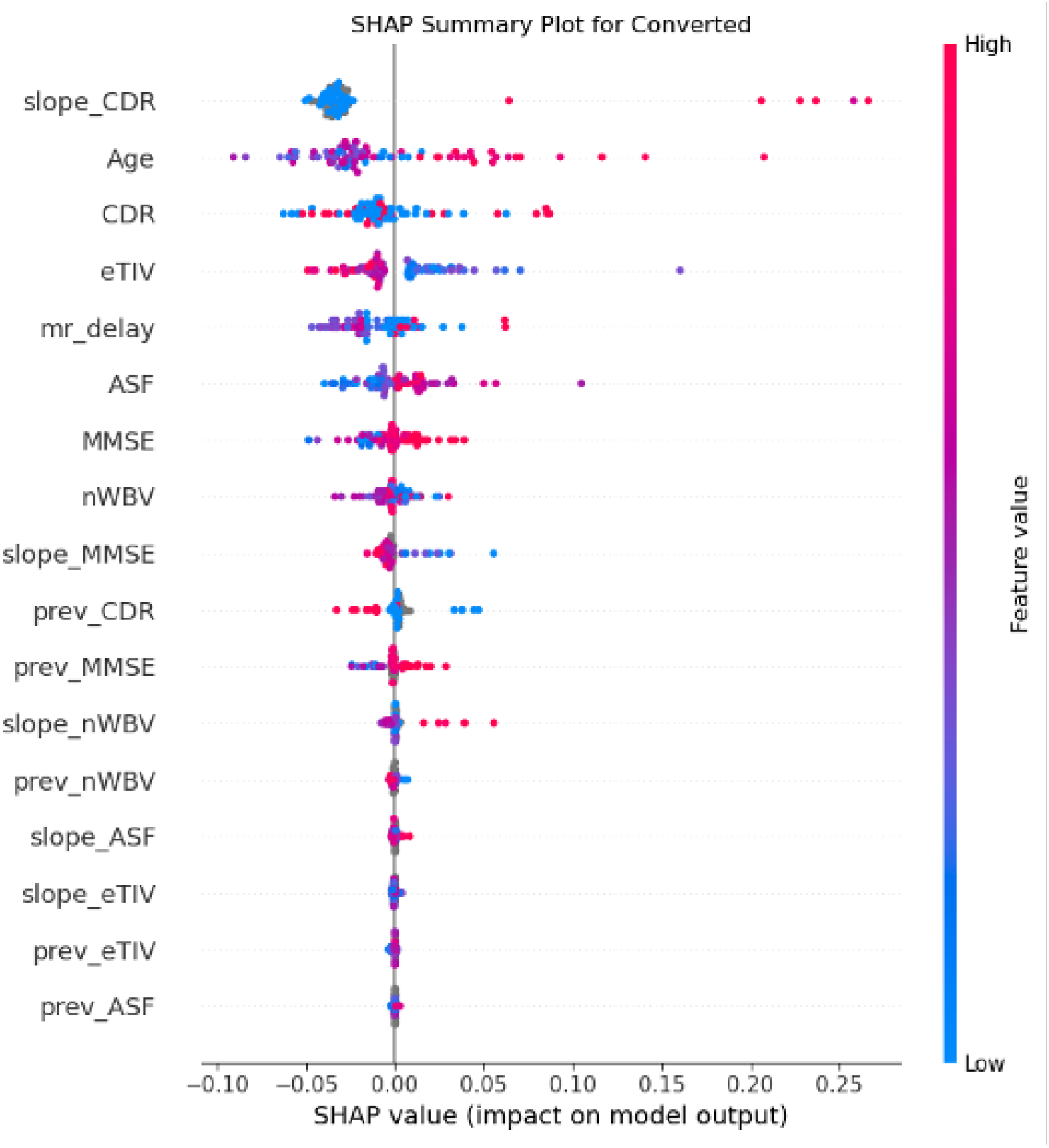
SHAP summary plot for the Converted class. Each point represents an observation, with SHAP values showing the direction and magnitude of each feature’s contribution to the model output. Higher values of slope CDR contributed most strongly to predictions for the Converted class, indicating the importance of longitudinal change in identifying transitional cases.

**Figure 9.**
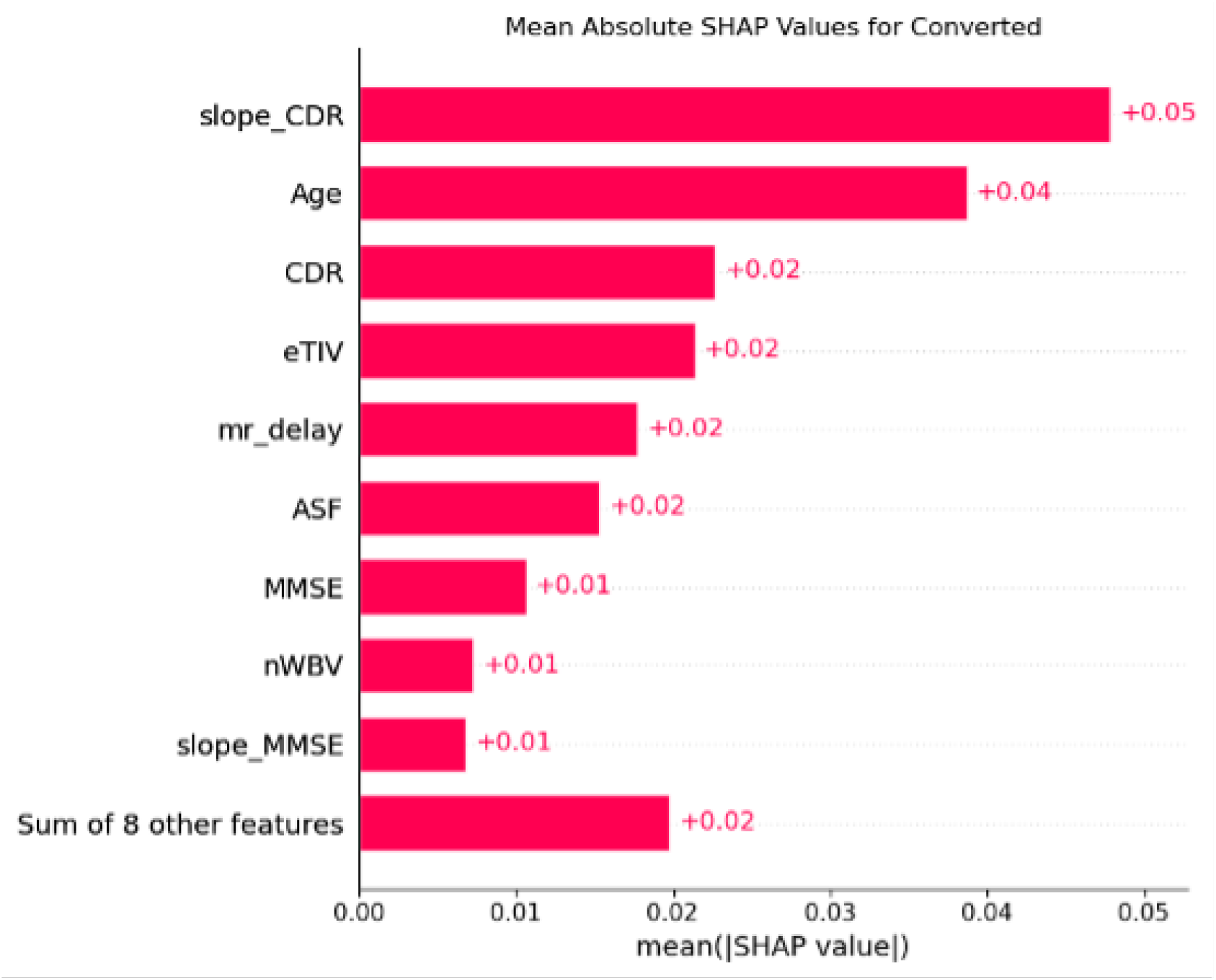
Mean absolute SHAP values for the Converted class. Slope CDR had the largest contribution, followed by Age and CDR.

For the Non-demented class, CDR was the dominant predictor. The mean absolute SHAP value for CDR was approximately 0.42, which was much larger than all other features. Age had the next highest contribution, approximately 0.03, followed by mr delay and eTIV, each around 0.02. Other variables, including ASF, prev_CDR, slope_CDR, MMSE, and nWBV, had smaller contributions of approximately 0.01. The SHAP summary plot in Figure 12 showed that lower CDR values increased the predicted probability of Non-demented classification, whereas higher CDR values reduced it. The mean absolute SHAP plot in Figure 13 further showed that Non-demented predictions were mainly driven by low dementia severity, with smaller contributions from Age, scan delay, and structural brain measures.

For the Demented class, CDR was also the strongest predictor, with a mean absolute SHAP value of approximately 0.41. Slope_CDR was the second most important feature, with a mean absolute SHAP value of approximately 0.04. Other features, including mr delay, prev_CDR, and MMSE, contributed approximately 0.02, while Age, eTIV, prev MMSE, and the remaining variables contributed less. The SHAP summary plot in Figure 10 showed that higher CDR values strongly increased the predicted probability of Demented classification, while lower CDR values reduced it. The mean absolute SHAP plot in Figure 11 confirmed that current dementia severity was the main driver of Demented classification.

**Figure 10.**
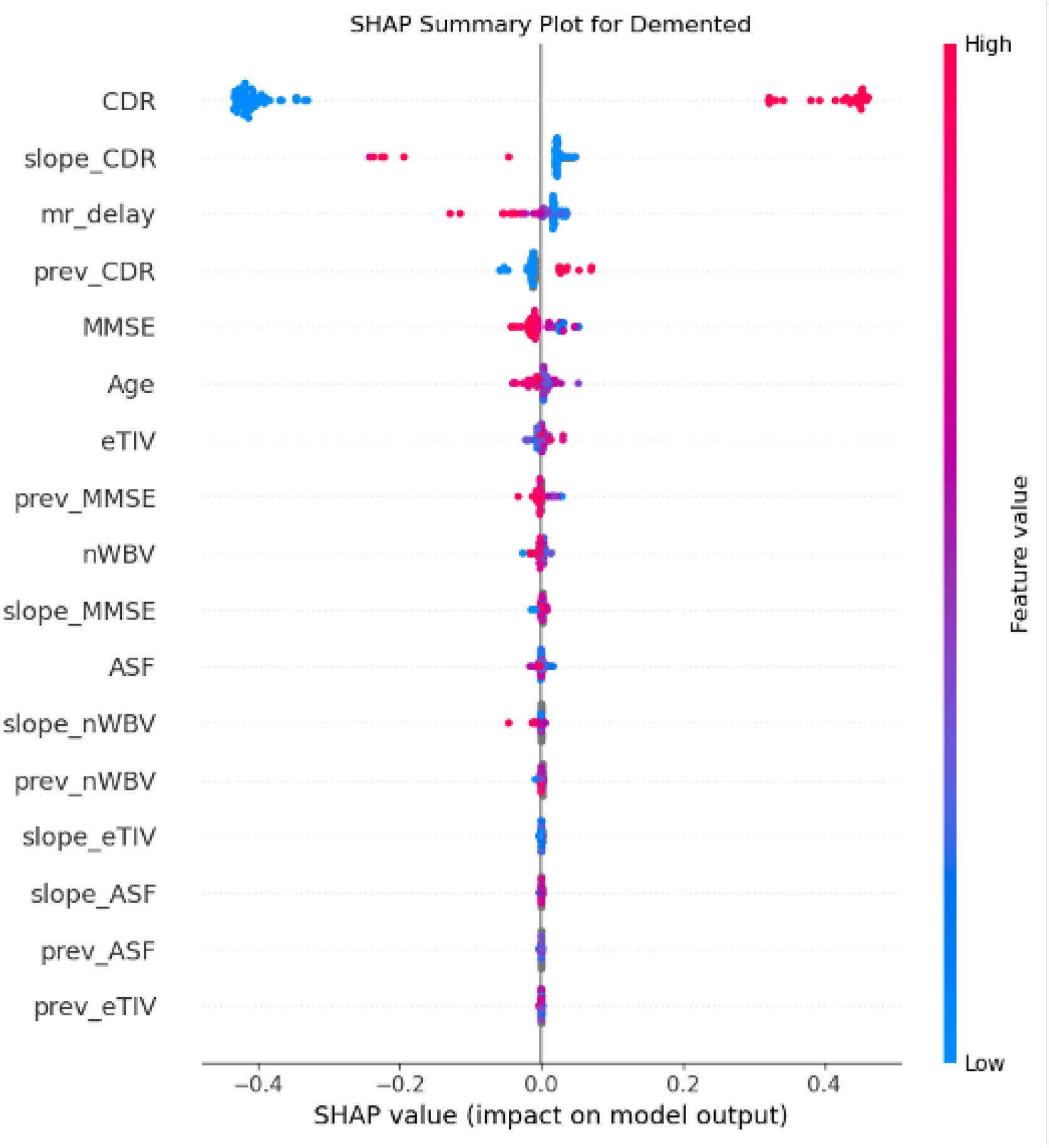
SHAP summary plot for the Demented class. CDR was the dominant predictor, with higher CDR values increasing the predicted probability of Demented classification. The contribution of slope CDR further shows that longitudinal change in clinical dementia rating influenced the model’s Demented-class predictions.

**Figure 11.**
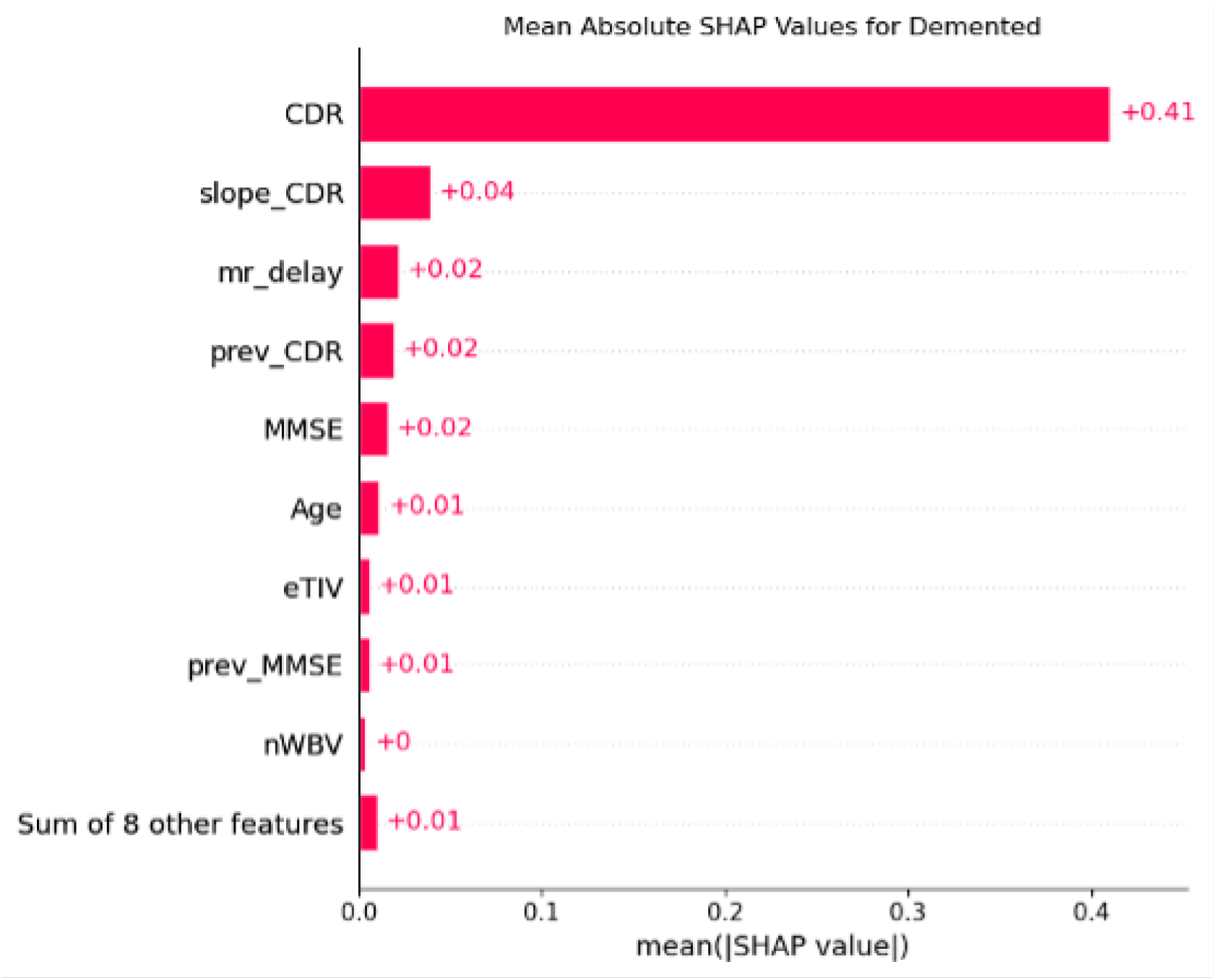
Mean absolute SHAP values for the Demented class. CDR had the largest contribution, showing that current dementia severity was the main driver of Demented classification.

**Figure 12.**
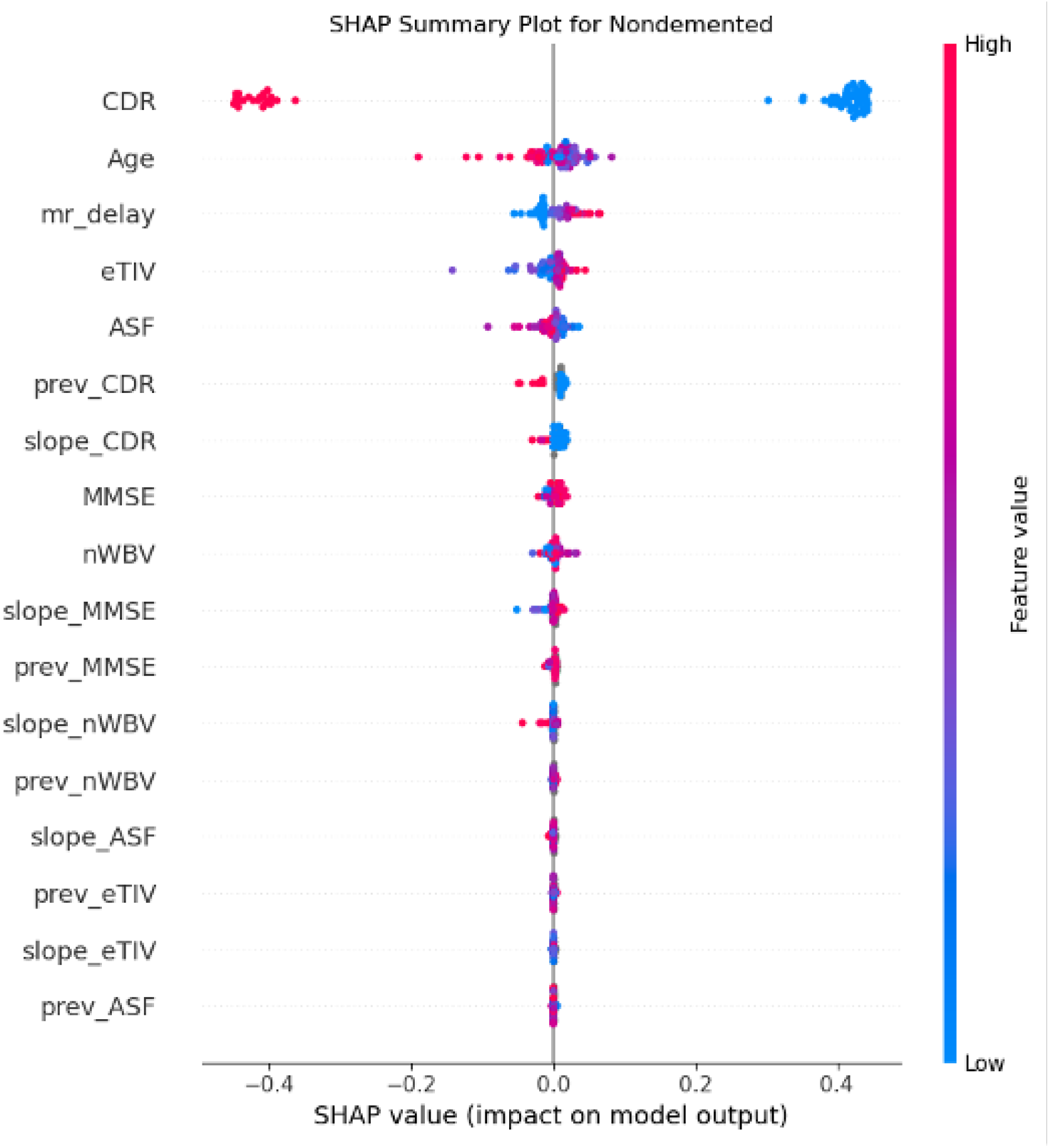
SHAP summary plot for the Non-demented class. Lower CDR values increased the predicted probability of Non-demented classification, while higher CDR values reduced the model output for this class. This pattern shows that CDR was central in separating Non-demented participants from those with dementia-related impairment.

**Figure 13.**
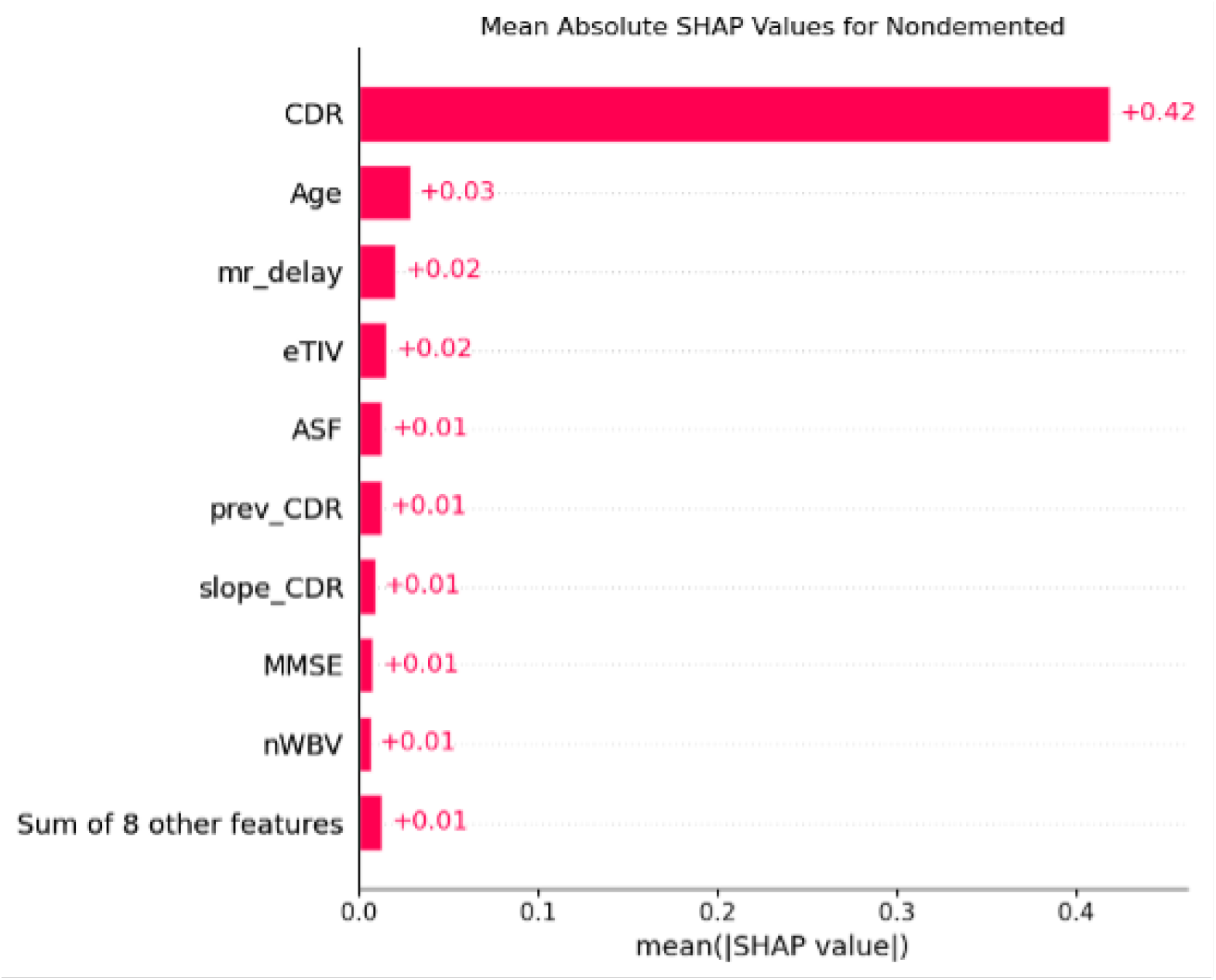
Mean absolute SHAP values for the Non-demented class. CDR had the largest contribution, indicating that low dementia severity was the main driver of Non-demented classification.

The SHAP results therefore show different feature patterns across the three diagnostic classes. For the Non-demented and Demented classifications, CDR was the dominant feature, indicating that current dementia severity was central to distinguishing between established diagnostic states. In contrast, for the Converted class, slope_CDR was the most important feature, suggesting that change in dementia severity over time was more useful for identifying transitional cases than a single current-visit measurement alone. This supports the main modeling result that adding temporal slope features improved classification performance.

Although MMSE had smaller SHAP contributions than CDR-related variables, it remains clinically important as a cognitive screening measure. A 1-point decline in MMSE may suggest mild cognitive worsening, but it should be interpreted cautiously because small changes can occur due to test–retest variability, fatigue, attention, education level, language differences, or the patient’s condition on the day of assessment. In clinical interpretation, a repeated decline of approximately 1–3 MMSE points over time is more meaningful, especially when it occurs together with worsening CDR or functional decline. Therefore, in this study, MMSE was interpreted as a supportive cognitive marker rather than a standalone diagnostic indicator.

Similarly, nWBV had a smaller SHAP contribution than CDR-related features, but it remains clinically relevant as a marker of brain atrophy. Lower nWBV reflects reduced normalized whole-brain volume; however, there is no single universal nWBV cutoff that defines clinically concerning brain-volume loss for all patients. The interpretation of nWBV depends on Age, baseline brain volume, follow-up duration, and rate of decline. Therefore, nWBV reduction becomes more concerning when it is progressive over time, greater than expected for Age, and accompanied by cognitive decline, increasing CDR, or conversion toward dementia.

Overall, the SHAP analysis suggests that the final model was clinically interpretable within this cohort. CDR was the strongest predictor for distinguishing Demented and Non-demented cases, while slope_CDR was most important for identifying Converted cases. MMSE and nWBV provided supportive information, but their clinical meaning was strongest when interpreted together with CDR, slope CDR, and longitudinal disease progression.

### 4.13. Learning Curve Analysis

The learning curve for the regularized model is summarized in Table 23 and shown in Figure 14. The training macro F1-score decreased as the training size increased. At 59 training observations, the training macro F1-score was 0.9948. At 297 training observations, it decreased to 0.8980. This indicates that the regularized model no longer maintained perfect training performance across all training sizes.

**Table 23.** Learning curve results for the regularized longitudinal Gradient boosting model.

| Training Size | Train F1 Mean | Train F1 SD | Validation F1 Mean | Validation F1 SD | Gap |
| --- | --- | --- | --- | --- | --- |
| 59 | 0.9948 | 0.0104 | 0.6446 | 0.0732 | 0.3503 |
| 118 | 0.9713 | 0.0225 | 0.7064 | 0.1563 | 0.2650 |
| 178 | 0.9530 | 0.0278 | 0.6639 | 0.1149 | 0.2890 |
| 237 | 0.8967 | 0.0306 | 0.6740 | 0.1022 | 0.2228 |
| 297 | 0.8980 | 0.0182 | 0.7212 | 0.0649 | 0.1768 |

**Figure 14.**
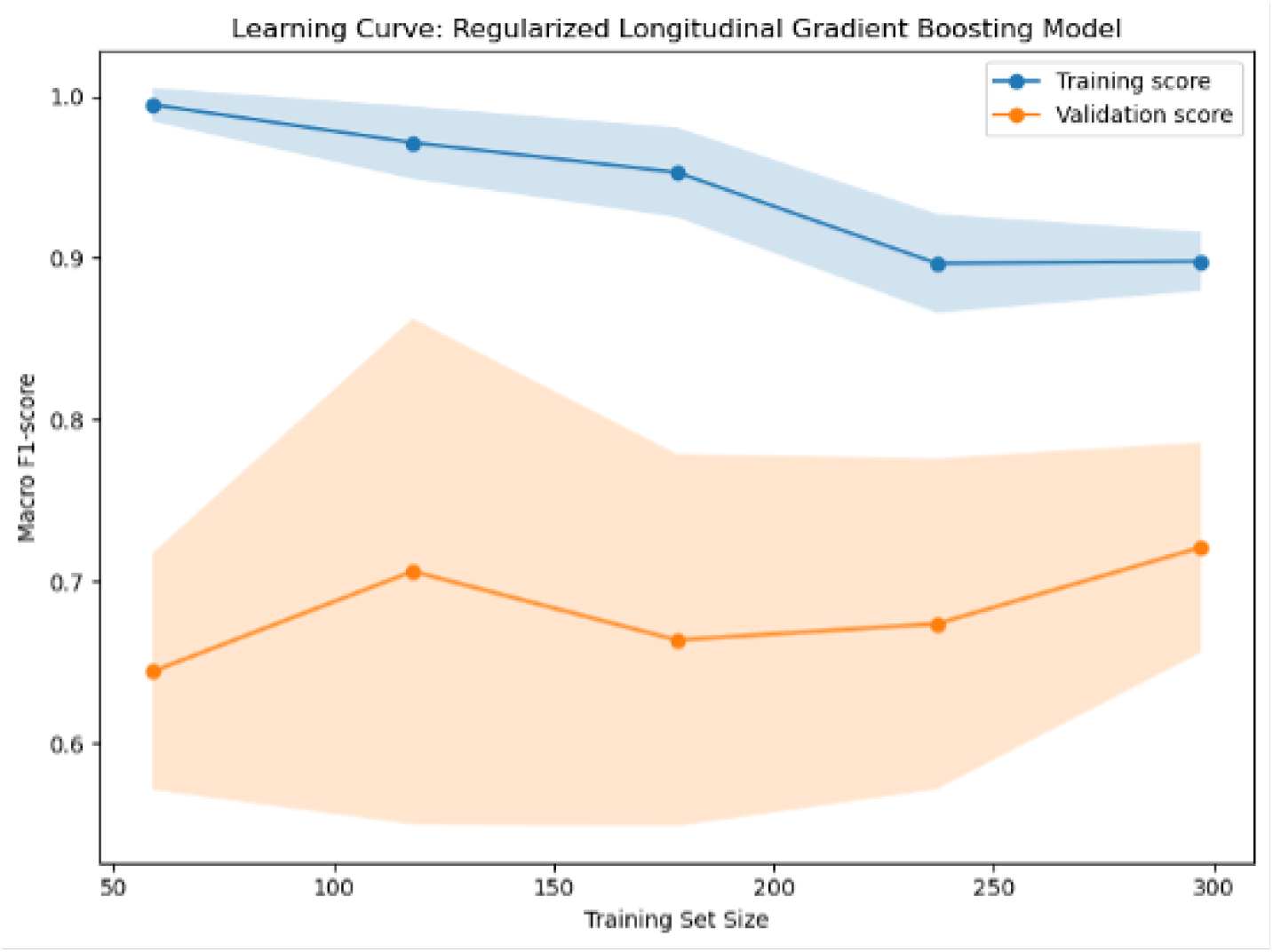
Learning curve for the regularized longitudinal Gradient boosting model. Training and validation macro F1-scores are shown across increasing training set sizes.

The validation macro F1-score increased overall from 0.6446 at 59 training observations to 0.7212 at 297 training observations. Although validation performance fluctuated at intermediate training sizes, the overall pattern suggested improved generalization with more training data. The generalization gap decreased from 0.3503 to 0.1768, indicating reduced evidence of overfitting as the training size increased. However, the remaining gap between training and validation performance suggests that some overfitting or sample-size limitation persisted.

### 4.14. Summary of Results

Overall, the original longitudinal Gradient boosting model achieved the highest held-out test performance, with an accuracy of 0.8816, macro F1-score of 0.7761, and weighted F1-score of 0.8604, as shown in Table 11. The regularized model reduced the perfect training fit observed in the original model, but its held-out test performance decreased to 0.8421 accuracy, 0.6815 macro F1-score, and 0.8060 weighted F1-score. Therefore, the original model was the best-performing model on the held-out test set, while the regularized model served as a sensitivity analysis for overfitting.

Across both models, the Demented and Non-demented classes were classified more accurately than the Converted class, as shown in Tables 12 and 13. The Converted class remained the most challenging group, as reflected by lower recall, lower F1-score, and lower AUC. Feature importance, permutation importance, and SHAP analysis consistently showed that CDR was the dominant predictor, while slope CDR provided additional longitudinal information, especially for identifying Converted cases. These findings suggest that temporal change features may improve dementia classification within this cohort, but prediction of transitional diagnostic states remains more difficult.

## 5. DISCUSSION

### 5.1. Comparison with Prior Work

Table 24 compares the present study with representative dementia machine learning and longitudinal modeling studies. The comparison includes studies based on ADNI, OASIS-2, MIRIAD, and neuropsychological test data, covering tasks such as dementia classification, MCI-to-AD conversion prediction, AD progression detection, and longitudinal disease progression modeling.

**Table 24.**
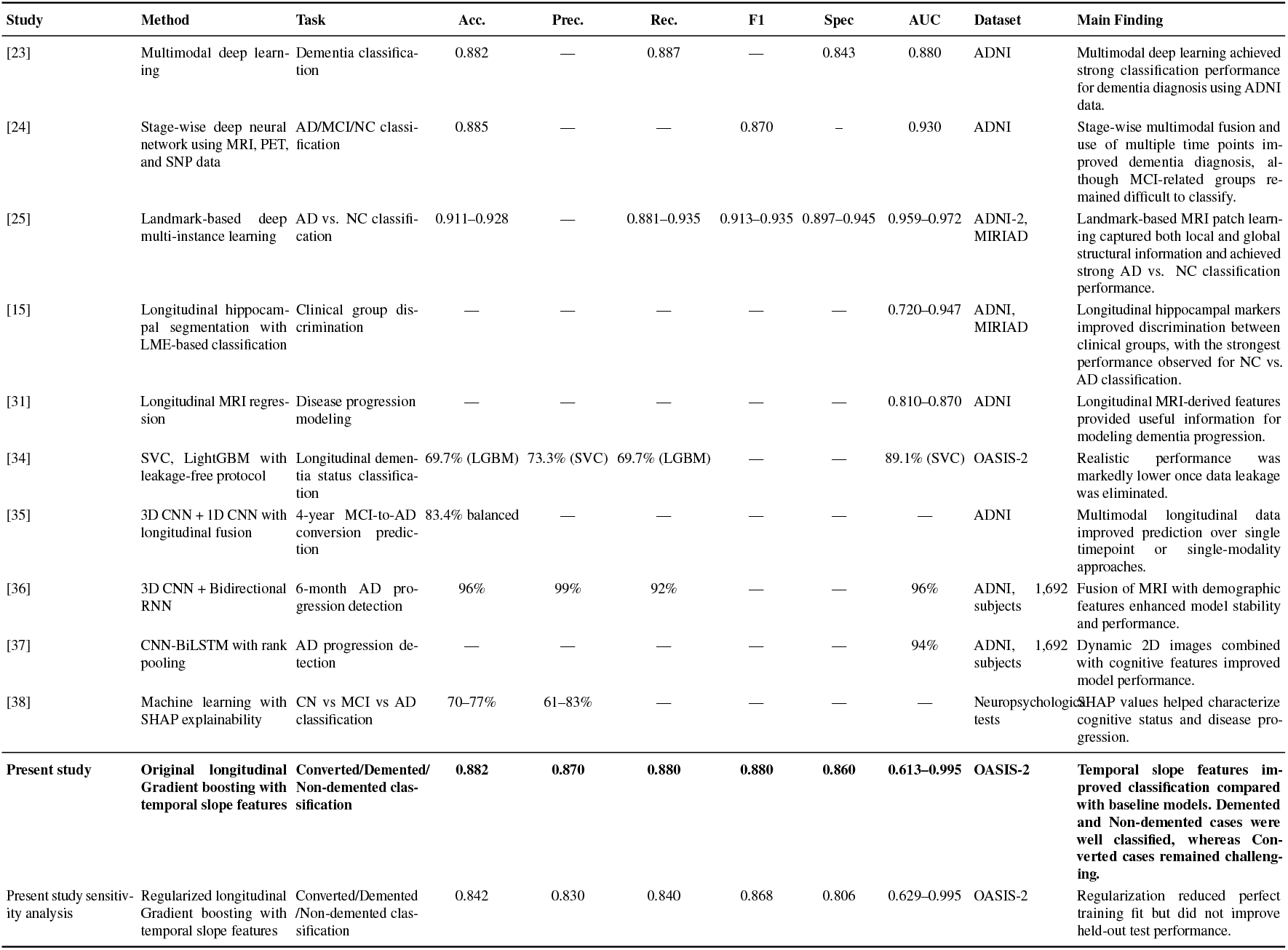
Comparative performance of the present study against representative machine learning and longitudinal classification studies in dementia. Acc. = accuracy; Prec. = precision; Rec. = recall/sensitivity; Spec. = specificity; F1 = F1-score; AUC = area under the ROC curve. “—” indicates that the metric was not reported. The bold row indicates the main proposed model in the present study.

The present study achieved an accuracy of 0.882, precision of 0.870, recall of 0.880, F1-score of 0.880, and class-level AUC values ranging from 0.613 to 0.995 on the OASIS-2 dataset. These results suggest promising predictive performance within this cohort. However, direct comparison with prior studies should be made cautiously because the studies differ in dataset, sample size, prediction task, imaging modality, validation strategy, feature construction, and reported evaluation metrics.

Several prior ADNI-based deep learning studies reported strong performance using multi-modal or longitudinal fusion approaches. For example, multimodal deep learning achieved an accuracy of 0.882, while a stage-wise deep neural network using MRI, PET, and SNP data achieved an accuracy of 0.885 and AUC of 0.930 [23, 24]. Landmark-based MRI patch learning also reported higher AD versus normal control classification performance, suggesting that local and global MRI structural information can support dementia classification [25].

The present study differs from many of these approaches by using a longitudinal gradient boosting model with explicitly engineered temporal slope features. This approach provides a simpler and more interpretable alternative to deep learning models. The leakage-free OASIS-2 study is particularly relevant because it showed that performance can decrease substantially when data leakage is carefully controlled [34]. This supports the need for patient-level splitting and cautious interpretation of model performance in longitudinal dementia datasets.

Progression-focused studies also show the value of longitudinal or multimodal information. Four-year MCI-to-AD conversion prediction using longitudinal neurocognitive tests and MRI data achieved balanced accuracy of 83.4%, while multimodal deep-learning fusion and recurrent architectures reported strong performance for AD progression detection [35–37]. Similarly, the present study found that slope-based temporal features improved classification compared with models using only current or previous visit measurements.

The class-specific AUC range in the present study was wide, indicating that performance was not uniform across diagnostic classes. Demented and Non-demented cases were classified more effectively, whereas Converted cases remained more challenging. This pattern is consistent with the difficulty of identifying transitional disease states, where individuals may share characteristics with both stable and impaired groups. Therefore, the findings should be interpreted as evidence that temporal features may improve classification within this cohort, rather than as proof of clinical readiness.

The use of SHAP-based explainability also connects the present study with prior explainable artificial intelligence work in dementia classification. Similar to previous work using SHAP to characterize cognitive status and disease progression, the present study used SHAP to identify clinically meaningful predictors, particularly CDR and slope CDR [38]. Overall, the comparison suggests that longitudinal temporal feature engineering combined with explainable machine learning may be useful for dementia classification, although future external validation is needed before broader clinical risk stratification.

## 6. LIMITATIONS AND FUTURE WORK

This study has several limitations. First, the analysis was based on the OASIS-2 longitudinal MRI dataset, which included 150 subjects and 373 observations. Although the dataset provides repeated measurements, the sample size is relatively small for machine learning model development and validation. This was especially important for the Converted class, which had fewer observations than the Demented and Non-demented classes. As a result, classification performance for Converted individuals was weaker, with lower recall, F1-score, and AUC values.

Second, the class distribution was imbalanced. The smaller number of Converted cases may have influenced the model toward better recognition of the more represented Demented and Non-demented groups. Although macro F1-score, weighted F1-score, class-specific metrics, and one-vs-rest AUC values were reported, the limited number of Converted cases makes interpretation of transitional-state classification less stable.

Third, the model was evaluated using a patient-level held-out test set and repeated stratified group cross-validation, but external validation was not performed. Therefore, the generalizability of the model to independent cohorts, different imaging protocols, and broader clinical populations remains uncertain. In addition, the original longitudinal Gradient boosting model achieved the best held-out test performance but showed perfect training performance, suggesting possible overfitting. Although a regularized model was evaluated as a sensitivity analysis, it did not improve held-out test performance.

Fourth, the temporal feature engineering approach relied on previous-visit values and slope-based rates of change. These features are useful for capturing progression, but they depend on the number and spacing of patient visits. Patients with fewer visits or irregular follow-up intervals may have less reliable slope estimates. The feature set was also limited to variables available in OASIS-2, including MMSE, CDR, eTIV, nWBV, ASF, Age, and MRI delay. More detailed biomarkers, such as hippocampal volume, cortical thickness, amyloid and tau measures, genetic factors, and vascular risk indicators, were not included.

Finally, the calibration analysis showed uncertainty in some probability bins, especially for the Converted class, partly because of small bin counts. Therefore, predicted probabilities should be interpreted cautiously for clinical risk stratification. Also, SHAP explanations improved interpretability but should not be interpreted as evidence of causality.

Future work should validate the proposed framework on larger and more diverse longitudinal dementia cohorts, including independent datasets such as ADNI or other multi-center studies. Further research should also incorporate richer neuroimaging, genetic, biomarker, and clinical risk variables to improve prediction of transitional cases. In addition, future models should explore more advanced longitudinal machine learning methods, including survival models, recurrent neural networks, and transformer-based temporal models. Finally, prospective clinical validation is needed to determine whether explainable longitudinal machine learning can support dementia screening, monitoring, and individualized risk stratification in real-world settings.

## 7. CONCLUSION

This study demonstrates the value of explainable longitudinal machine learning for modeling dementia progression using cognitive and MRI-derived features. By incorporating current measurements, previous-visit information, and slope-based temporal changes, the proposed longitudinal Gradient boosting framework captured both cross-sectional differences and progression-related patterns in the OASIS-2 cohort.

The original longitudinal Gradient boosting model achieved the best held-out test performance, outperforming the baseline models in accuracy, macro F1-score, and weighted F1-score. The model showed stronger classification performance for the Demented and Non-demented groups, while the Converted class remained the most difficult to identify. This highlights the challenge of predicting transitional diagnostic states, where individuals may show overlapping cognitive and structural characteristics between stable normal aging and established dementia.

Feature importance, permutation importance, and SHAP-based explanations showed that CDR was the strongest predictor of dementia classification, while slope-based CDR contributed additional longitudinal information. Cognitive variables were more influential than global brain-volume measures, although MRI-derived variables such as nWBV, eTIV, and ASF still provided supportive predictive information. These findings suggest that combining clinical severity measures with temporal change features may improve dementia progression modeling.

Although the model achieved promising performance within this cohort, its findings should be interpreted with caution due to the small sample size, class imbalance, and lack of external validation. Future work should evaluate the framework in larger, more diverse longitudinal cohorts, incorporate richer biomarkers such as regional brain volumes, cortical thickness, amyloid and tau imaging, genetic markers, and vascular risk factors, and explore advanced temporal modeling approaches. Prospective validation will also be necessary to determine whether explainable longitudinal machine learning can support dementia screening, monitoring, and individualized risk stratification in real-world clinical settings.

## Data Availability

The data that support the findings of this study are publicly available and can be accessed from the OASIS longitudinal dataset \href{https://www.oasis-brains.org}{https://www.oasis-brains.org

## Clinical trial number

Not applicable

## Ethics approval and consent to participate

This retrospective study used aggregated, secondary data obtained from the publicly available OASIS longitudinal dataset. We confirm that no personally identifiable or confidential information was accessed or used at any stage of the analysis.

## Consent for publication

Not applicable

## Data availability statement

The data that support the findings of this study are publicly available and can be accessed from the OASIS longitudinal dataset https://www.oasis-brains.org

## Author contributions

**Gifty Duah:** Conceptualization, Methodology, Data curation, Visualization, Project Administration, Resources, Formal Analysis, Software, Writing-Original draft, Writing-Reviewing and Editing, Investigation and Validation. **Eric Nyarko:** Conceptualization, Methodology, Writing-Original draft, Writing-Reviewing and Editing and Validation. **Justice Yaw Effah:** Writing-Reviewing and Editing, Visualization. **Isaac Boateng Numoah:** Writing-Reviewing and Editing, Visualization and Validation. **Anani Lotsi:** Conceptualization, Writing-Reviewing and Editing, Visualization and Validation.

## Acknowledgements

We would like to acknowledge and thank OASIS for making the anonymized secondary data available for analysis.

## References

[1] Fatih Gelir et al. Heterogeneity in cognitive decline trajectories across Alzheimer’s disease and related dementias. Alzheimer’s & Dementia, 21(1):e14501, 2025. doi: 10.1002/alz.14501.

[2] Martin Prince, Renata Bryce, Emiliano Albanese, Anders Wimo, Wagner Ribeiro, and Cleusa P. Ferri. The global prevalence of dementia: A systematic review and metaanalysis. Alzheimer’s & Dementia, 9(1):63–75, 2013. doi: 10.1016/j.jalz.2012.11.007.

[3] World Health Organization. Global status report on the public health response to dementia. Technical report, World Health Organization, 2021. URL https://www.who.int/publications/i/item/9789240033245.

[4] Alzheimer’s Association. 2023 Alzheimer’s disease facts and figures. Alzheimer’s & Dementia, 19(4):1598–1695, 2023. doi: 10.1002/alz.13016.

[5] Gill Livingston, Jonathan Huntley, Andrew Sommerlad, David Ames, Clive Ballard, Sube Banerjee, Carol Brayne, Alistair Burns, Jiska Cohen-Mansfield, Claudia Cooper, et al. Dementia prevention, intervention, and care: 2020 report of the Lancet Commission. The Lancet, 396(10248):413–446, 2020. doi: 10.1016/S0140-6736(20)30367-6.

[6] Reisa A. Sperling, Paul S. Aisen, Laurel A. Beckett, David A. Bennett, Suzanne Craft, Anne M. Fagan, Takeshi Iwatsubo, Clifford R. Jack, Jeffrey Kaye, Thomas J. Montine, et al. Toward defining the preclinical stages of Alzheimer’s disease: Recommendations from the National Institute on Aging–Alzheimer’s Association workgroups. Alzheimer’s & Dementia, 7(3):280–292, 2011. doi: 10.1016/j.jalz.2011.03.003.

[7] Anders Wimo, Maëlenn Guerchet, Gemma-Claire Ali, Yu-Tzu Wu, A. Matthew Prina, Bengt Winblad, Linus Jönsson, Zhaorui Liu, and Martin Prince. The worldwide costs of dementia 2015 and comparisons with 2010. Alzheimer’s & Dementia, 13(1):1–7, 2017. doi: 10.1016/j.jalz.2016.07.150.

[8] Lars Lau Rakêt. Statistical separation of genetic and environmental effects in longitudinal data: design and analysis of longitudinal twin studies. Annals of Applied Statistics, 14(2):949–975, 2020. doi: 10.1214/19-AOAS1310.

[9] Clifford R. Jack, David S. Knopman, William J. Jagust, Leslie M. Shaw, Paul S. Aisen, Michael W. Weiner, Ronald C. Petersen, and John Q. Trojanowski. Hypothetical model of dynamic biomarkers of the Alzheimer’s pathological cascade. The Lancet Neurology, 9(1):119–128, 2010. doi: 10.1016/S1474-4422(09)70299-6.

[10] Marshal F. Folstein, Susan E. Folstein, and Paul R. McHugh. “Mini-Mental State”: A practical method for grading the cognitive state of patients for the clinician. Journal of Psychiatric Research, 12(3):189–198, 1975. doi: 10.1016/0022-3956(75)90026-6.

[11] John C. Morris. The Clinical Dementia Rating (CDR): Current version and scoring rules. Neurology, 43(11):2412–2414, 1993. doi: 10.1212/WNL.43.11.2412.

[12] Ronald C. Petersen. Mild cognitive impairment as a diagnostic entity. Journal of Internal Medicine, 256(3):183–194, 2004. doi: 10.1111/j.1365-2796.2004.01388.x.

[13] Ronald C. Petersen, J. C. Stevens, Mary Ganguli, Eric G. Tangalos, Jeffrey L. Cummings, and Steven T. DeKosky. Practice parameter: Early detection of dementia: Mild cognitive impairment (an evidence-based review). Neurology, 56(9):1133–1142, 2001. doi: 10.1212/WNL.56.9.1133.

[14] Ahmed A. Moustafa, Doaa H. Hewedi, Abeer M. Eissa, Dorota Frydecka, and Blazej Misiak. Cognitive decline in normal aging and its prevention: a review on non-pharmacological lifestyle strategies. Current Aging Science, 13(3):199–215, 2020. doi: 10.2174/1874609812666191202133314.

[15] Carlos Platero. Longitudinal neuroimaging hippocampal markers for diagnosing Alzheimer’s disease. Neuroinformatics, 20(1):85–100, 2022. doi: 10.1007/s12021-021-09521-1.

[16] Evan Fletcher, Mekala Raman, Peg Huebner, Audrey Liu, Dan Mungas, Owen Carmichael, and Charles DeCarli. Loss of fornix white matter volume as a predictor of cognitive impairment in cognitively normal elderly individuals. JAMA Neurology, 70(11):1389–1395, 2013. doi: 10.1001/jamaneurol.2013.3263.

[17] Clifford R. Jack, Ronald C. Petersen, Yue Cheng Xu, Peter C. O’Brien, Glenn E. Smith, Robert J. Ivnik, Bradley F. Boeve, Stephen C. Waring, Eric G. Tangalos, and Emre Kokmen. Prediction of AD with MRI-based hippocampal volume in mild cognitive impairment. Neurology, 52(7):1397–1403, 1999. doi: 10.1212/WNL.52.7.1397.

[18] Nick C. Fox and Peter A. Freeborough. Brain atrophy progression measured from registered serial MRI: validation and application to Alzheimer’s disease. Journal of Magnetic Resonance Imaging, 7(6):1069–1075, 1997. doi: 10.1002/jmri.1880070614.

[19] Brandon E. Gavett, Keith F. Widaman, Dan Mungas, Paul K. Crane, David A. Bennett, and Mary N. Tomboulouand. Alzheimer’s pathology explains relationship between dementia severity and cognitive reserve. Alzheimer’s & Dementia, 17(1):e050429, 2021. doi: 10.1002/alz.050429.

[20] Raymond Y. Lo, Alan E. Hubbard, Leslie M. Shaw, John Q. Trojanowski, Ronald C. Petersen, Paul S. Aisen, Michael W. Weiner, and William J. Jagust. Longitudinal change of biomarkers in cognitive decline. Archives of Neurology, 68(10):1257–1266, 2011. doi: 10.1001/archneurol.2011.123.

[21] Heiko Braak and Eva Braak. Neuropathological stageing of Alzheimer-related changes. Acta Neuropathologica, 82(4):239–259, 1991. doi: 10.1007/BF00308809.

[22] Patrizia Ribino et al. Multivariate longitudinal clustering for dementia subgroup identification. Journal of Biomedical Informatics, 161:104752, 2025. doi: 10.1016/j.jbi.2024.104752.

[23] Fan Zhang, Zhiwei Li, Baiying Zhang, Haoran Du, Bo Wang, and Xiaodong Zhang. Multi-modal deep learning model for auxiliary diagnosis of Alzheimer’s disease. Neurocomputing, 361:185–195, 2019. doi: 10.1016/j.neucom.2019.04.093.

[24] Tao Zhou, Kim-Han Thung, Xiaofeng Zhu, and Dinggang Shen. Effective feature learning and fusion of multimodality data using stage-wise deep neural network for dementia diagnosis. Human Brain Mapping, 40(3):1001–1016, 2019. doi: 10.1002/hbm.24428.

[25] Mingxia Liu, Jun Zhang, Ehsan Adeli, and Dinggang Shen. Landmark-based deep multi-instance learning for brain disease diagnosis. Medical Image Analysis, 43: 157–168, 2018. doi: 10.1016/j.media.2017.10.005.

[26] Scott M. Lundberg and Su-In Lee. A unified approach to interpreting model predictions. In Advances in Neural Information Processing Systems, volume 30, pages 4765–4774, 2017. URL https://proceedings.neurips.cc/paper/2017/hash/8a20a8621978632d76c43dfd28b67767-Abstract.html.

[27] Scott M. Lundberg, Gabriel Erion, Hugh Chen, Alex DeGrave, Jordan M. Prutkin, Bala Nair, Ronit Katz, Jonathan Himmelfarb, Nisha Bansal, and Su-In Lee. From local explanations to global understanding with explainable AI for trees. Nature Machine Intelligence, 2(1):56–67, 2020. doi: 10.1038/s42256-019-0138-9.

[28] Erico Tjoa and Cuntai Guan. A survey on explainable artificial intelligence (XAI): Toward medical XAI. IEEE Transactions on Neural Networks and Learning Systems, 32(11):4793–4813, 2021. doi: 10.1109/TNNLS.2020.3027314.

[29] Daniel S. Marcus, Anthony F. Fotenos, John G. Csernansky, John C. Morris, and Randy L. Buckner. Open access series of imaging studies: Longitudinal MRI data in nondemented and demented older adults. Journal of Cognitive Neuroscience, 22(12): 2677–2684, 2010. doi: 10.1162/jocn.2009.21407.

[30] Dan Mungas, Bruce R. Reed, William J. Jagust, Charles DeCarli, Wendy J. Mack, Joel H. Kramer, Michael W. Weiner, Norbert Schuff, and Helena C. Chui. MRI predictors of cognition in subcortical ischemic vascular disease and Alzheimer’s disease. Neurology, 57(12):2229–2235, 2001. doi: 10.1212/WNL.57.12.2229.

[31] Gengsheng Chen, B. Douglas Ward, Chunming Xie, Wenjun Li, Gang Chen, Joseph S. Goveas, Piero G. Antuono, and Shi-Jiang Li. Staging Alzheimer’s disease risk by sequencing brain function and structure, cerebrospinal fluid, and cognition biomarkers. Journal of Alzheimer’s Disease, 54(3):983–993, 2016. doi: 10.3233/JAD-160537.

[32] Fabian Pedregosa, Gaël Varoquaux, Alexandre Gramfort, Vincent Michel, Bertrand Thirion, Olivier Grisel, Mathieu Blondel, Peter Prettenhofer, Ron Weiss, Vincent Dubourg, et al. Scikit-learn: Machine learning in Python. Journal of Machine Learning Research, 12:2825–2830, 2011. URL https://jmlr.csail.mit.edu/papers/v12/pedregosa11a.html.

[33] Sid E. O’Bryant, Stephen C. Waring, C. Munro Cullum, James Hall, Laura Lacritz, Paul J. Massman, Philip J. Lupo, Joan S. Reisch, and Rachelle Doody. Staging dementia using Clinical Dementia Rating Scale Sum of Boxes scores: A Texas Alzheimer’s research consortium study. Archives of Neurology, 65(8):1091–1095, 2008. doi: 10.1001/archneur.65.8.1091.

[34] Mohammad Mahdi Ghiasi, R. Falck, T. Liu-Ambrose, and Roger C. Tam. Explainable machine learning for predicting longitudinal dementia status: Establishing a leakage-free benchmark. PLOS Digital Health, 2026.

[35] Rohan Bapat, D. Ma, and Tim Q. Duong. Predicting four-year’s alzheimer’s onset using longitudinal neurocognitive tests and mri data using explainable deep convolutional neural networks. Journal of Alzheimer’s Disease, 2023.

[36] Nasir Rahim, Shaker El-Sappagh, Sajid Ali, Khan Muhammad, Javier Del Ser, and Tamer Abuhmed. Prediction of alzheimer’s progression based on multimodal deep-learning-based fusion and visual explainability of time-series data. Information Fusion, 2022.

[37] Nasir Rahim, T. Abuhmed, S. Mirjalili, Shaker El-Sappagh, and Khan Muhammad. Time-series visual explainability for alzheimer’s disease progression detection for smart healthcare. Alexandria Engineering Journal, 2023.

[38] Lombardi, D. Diacono, N. Amoroso, P. Biecek, A. Monaco, L. Bellantuono, E. Pantaleo, G. Logroscino, R. Blasi, S. Tangaro, and R. Bellotti. A robust framework to investigate the reliability and stability of explainable artificial intelligence markers of mild cognitive impairment and alzheimer’s disease. Brain Informatics, 2022.

